# Identification of Causal Pathways among Gut Microbiota, Food Intake and Telomere Length: A Mendelian Randomization Study

**DOI:** 10.1101/2022.09.22.22280255

**Authors:** Lei Hou, Yuanyuan Yu, Chuandi Jin, Lei Zhang, Yilei Ge, Xinhui Liu, Sijia Wu, Fengtong Qian, Yutong Wu, Yifan Yu, Qingxin Luo, Yina He, Yue Feng, Yun Wei, Hongkai Li, Fuzhong Xue

## Abstract

**Background:** Dietary habit plays an important role in the composition and function of gut microbiota which possibly manipulates host eating behavior. Gut microflora and nutritional imbalance are associated with telomere length (TL). However, the causality among them remains unclear.

**Methods:** Firstly, we calculate the significance threshold based on genetic correlations. Then we perform bi-directional Mendelian Randomization (MR) analyses among 82 food intakes (FIs) (UK Biobank, N=455,146), 95 gut microbial traits (Flemish Gut Flora Project, N=2,223) and TL (genome-wide meta-analysis from 15 cohorts, N=37,684) using summary-level data from large genome-wide association studies. Fixed-effect inverse variance weighting is the main analysis method and the other eight two-sample MR methods and three sensitivity analyses are performed. Finally, GO enrichment analyses are used to investigate the bio-function.

**Results:** Several bi-directional causal relationships among gut microbiota, FIs and TL are obtained by two-sample MR. Overall, we find suggestive evidence of three main causal pathways among them. Drinking more glasses of water per day is able to affect the habit of eating dried fruit through the host gut microbiota (Barnesiella). The change of one gut microbiota taxon (Collinsella) in the host causally influences another gut microbiota taxon (Lactonccus) through the diet habits (intake of oil-based spread). Additionally, the TL alters the habits of drinking ground coffee and further affects the gut microbiota (Acidaminococcaceae). GO enrichment analysis further confirmed the MR results.

**Conclusion:** TL has an impact on diet habits and gut microbiota and there are bi-directional relationships between diet habits and gut microbiota.

## 1 Introduction

In human growth and development, the composition and function of gut microbiota are physiologically influenced by varieties of environmental factors, among which dietary habit plays an important role[1-3]. Inversely, the preferences of the host may be influenced by the gut microbiota[4-6]. Also, gut microbiota possibly manipulates host eating behaviors in ways that promote their fitness at the expense of host fitness because of evolutionary conflict between host and microbes in the gut[7]. We wonder if there is a feedback loop existed between the preferences of the host for a particular dietary and the composition of gut microbiota in host. But the causality between changes in the microbiota composition and host feeding behavior remains unclear in observational studies. Adherence to a Western dietary pattern or a “prudent” pattern (based on high factor loadings of fruits, vegetables, nuts, fish, chicken and turkey without skin) was found to be associated with measures of β-diversity, but not α-diversity for 517 older men [8]. Claesson et al. gathered food frequency questionnaires (FFQ) data on 178 older men and women in Ireland and found that dietary diversity was significantly associated with improved health parameters, with the grouping “low fat/high fibre” considered to have the most diverse diet and microbial profile[9]. Bolte et al.[10] found that dietary patterns were consistently correlated with groups of bacteria with shared functional roles in both, health and disease, based on 173 dietary factors and the microbiome of 1425 individuals spanning four cohorts. However, there are a limited number of studies investigating the changes in long-term habitual diet in response to microbial composition[11-14]. For example, a 2016 study revealed that gut bacterial proteins could influence host’s control of appetite dependent on the bacterial growth phase[11]. Further studies of diet–microbiome relations remain required.

Telomeres are sections of repetitive DNA located at the ends of chromosomes and play an essential role in maintaining chromosome stability[15]. Loss of telomeres occurs after each cell division due to the inability of DNA polymerase to completely replicate chromosomal ends. Evidences from previous studies suggested a potential link between telomere attrition and microbes[16-21]. Maeda et al.[17] found that the length of the combined range of telomere and the methylated sub-telomere was correlated with the increase of bacteria species and the numerical superiority of certain strains in feces. Additionally, obesity, which is strongly influenced by the microbiota, exacerbates telomere shortening in humans[16,21]. A five-year longitudinal study investigating how diet impacts telomere length (TL) in humans showed a strong correlation between the effects of a Mediterranean diet with anti-inflammatory properties and a delay in telomere shortening[18]. A systematic review including 17 studies revealed that dietary patterns and different food groups were assessed in relation to TL. Mediterranean dietary pattern was related to longer TL and eating fruits or vegetables was positively associated with TL, but a reverse association between TL and intake of cereals, processed meat, and fats and oils was reported[22]. Therefore, a dysbiotic gut microbiota as a result of aging or through nutritional imbalance could indirectly lead to telomere attrition[19,20].

Mendelian Randomization (MR) can be used to assess the causal effect of an exposure on an outcome using genetic variants as instrumental variables (IVs). Three assumptions of MR should be satisfied: 1) relevance: the genetic variants are associated with exposure; 2) exchangeability: the genetic variants are not associated with any confounders of exposure and outcome; 3) exclusion restriction: the genetic variants are conditionally independent of the outcome given the exposure and confounders. It overcomes the difficulties of residual confounding and reverse causation. Recent genetic studies have demonstrated that the host genetic variants influence the gut microbiota composition [23-25]. A phenome□wide association study revealed that eleven single nucleotide polymorphisms (SNPs) were significantly associated with food intakes (FIs) after Bonferroni correction [26]. The locus on LCT, which was found to be associated with multiple microbial taxa, seemed modulated by lactose intake[27]. The MR studies related to TL mainly focus on the causal relationship between TL and behavioral habits [28] and aging□related outcomes[29,30]. However, there is no MR literature among TL, FIs and gut microbiota. The causality among changes in the microbiota composition, host feeding behaviors and TL remains to be established.

## 2 Methods

### 2.1 Study design overview

The design of this study is shown in Figure 1, we firstly filter the host genetic variants as IVs to explore the causal relationships among gut microbiota, FIs and TL. We perform a large-scale bi-directional MR analyses among 95□gut microbial traits, 82 kinds of FIs and TL using Genome-Wide Association Study (GWAS) summary data. The significance threshold is calculated by clustering the genetic correlations using the high-definition likelihood (HDL) method. Next, we construct a causal network structure among FI, gut microbiota and TL. Finally, GO enrichment analyses are used to investigate the bio-function.

**Figure 1.**
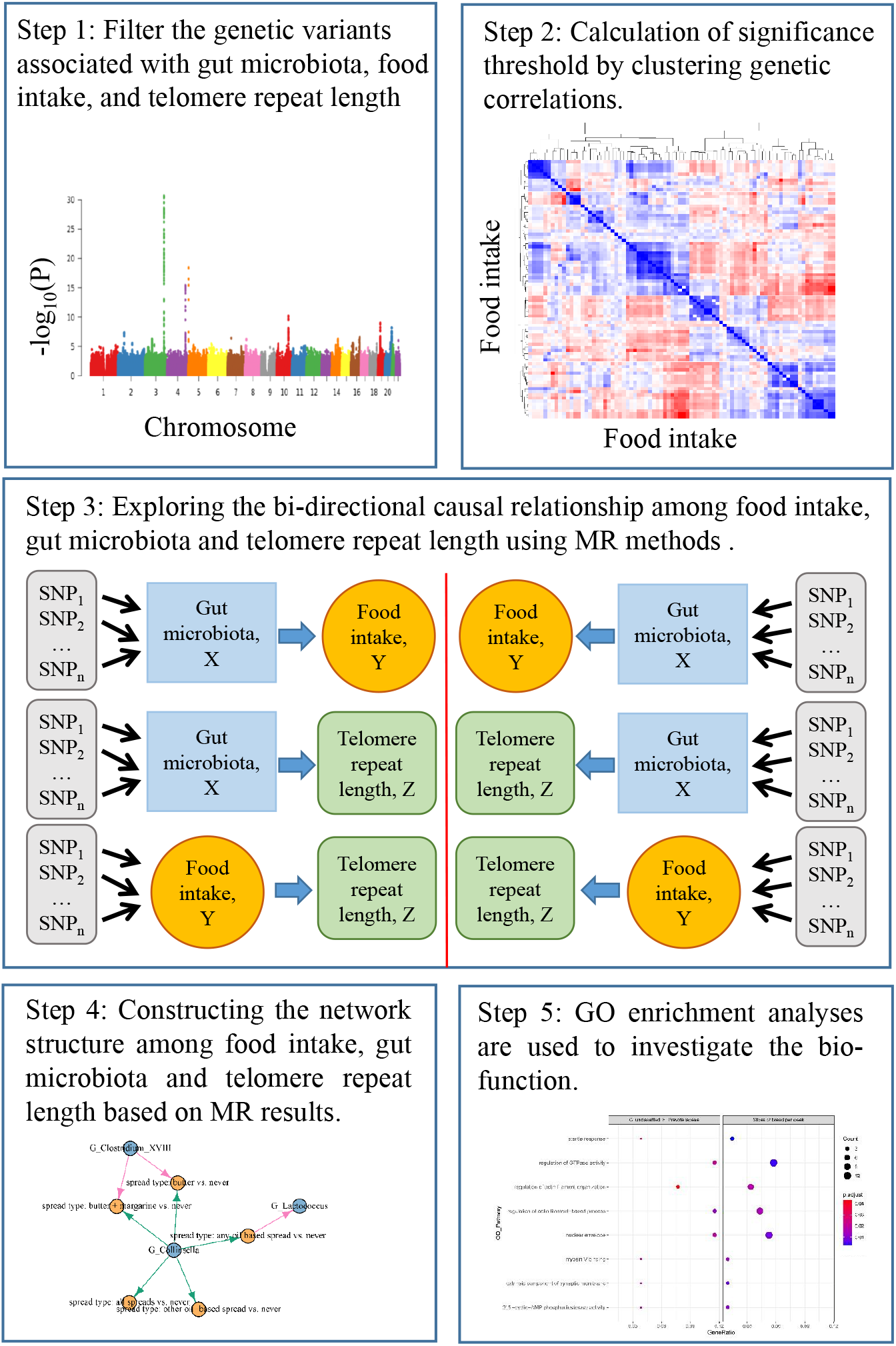
The design of this study. (Step1) We firstly filter the genetic variants associated with gut microbiota, food intakes, and telomere length (TL). (Step2) We calculate the significance threshold by clustering genetic correlations among them using the high-definition likelihood (HDL) method. (Step3) We explore the bi-directional causal relationships among them using MR analysis. (Step4) We construct the network structure among them. (Step5) GO enrichment analyses are used to investigate the bio-function.

### 2.2 Data sources and instruments selection

#### Gut microbiota

We leverage summary statistics from a GWAS of gut microbiota conducted among the Flemish Gut Flora Project (FGFP, n=2223) using 16S rRNA gene sequences and host genotype data[31]. After rank normal transformation (RNT), 95□continuous traits are carried forward to GWAS analyses as microbial traits (Supp. Table 1). We select SNPs at a threshold for genome-wide significance (P < 1 × 10^−5^) from this GWAS as genetic instruments. Then we prune the variants by linkage disequilibrium (LD) (r^2^ > 0.01, clumping window = 1000 kbp) and minimum allele frequency (MAF) (MAF < 5%). Mean F statistics [32] of all instrumental variants of gut microbiota are above 10.

#### Food intake

The GWAS summary statistics datasets for FIs in this study are from the UK Biobank [33]. We use summary data of 82 FIs from FFQ in UK Biobank (Supp. Table 2) [34]. The UKB FFQ consists of quantitative continuous variables, ordinal non-quantitative variables depending on overall daily/weekly frequency, food types, or foods never eaten. Ordinal variables are ranked and set to quantitative values, while food types or foods never eaten are converted into a series of binary variables. Corresponding FFQ question for each dietary habit can be looked up in the UKB Data Showcase (http://biobank.ndph.ox.ac.uk/showcase/). All 82 FIs are then adjusted for age in months and sex, followed by inverse RNT FIs. We select SNPs at a threshold for suggestive genome-wide significance (P < 5 × 10^−5^) from the GWAS for each type of FIs. Quality control of LD is the same as the gut microbiota.

#### Telomere length

Codd et al. [35] reported a genome-wide meta-analysis of 37,684 individuals from 15 cohorts, along with an additional 10,739 individuals from 6 cohorts for replication of selected variants. Mean leukocyte TL was measured using a quantitative PCR–based technique [36,37], which expresses TL as a ratio (T/S) of telomere repeat length (T) to copy the number of a single copy gene (S). The standard of instrumental variants selection is the same as the FIs.

Description of GWAS summary data for gut microbiota, FIs and TL can be seen in Table 1. Ethical approval for each study included in this article can be found in the original articles [31,34,35].

**Table 1.**
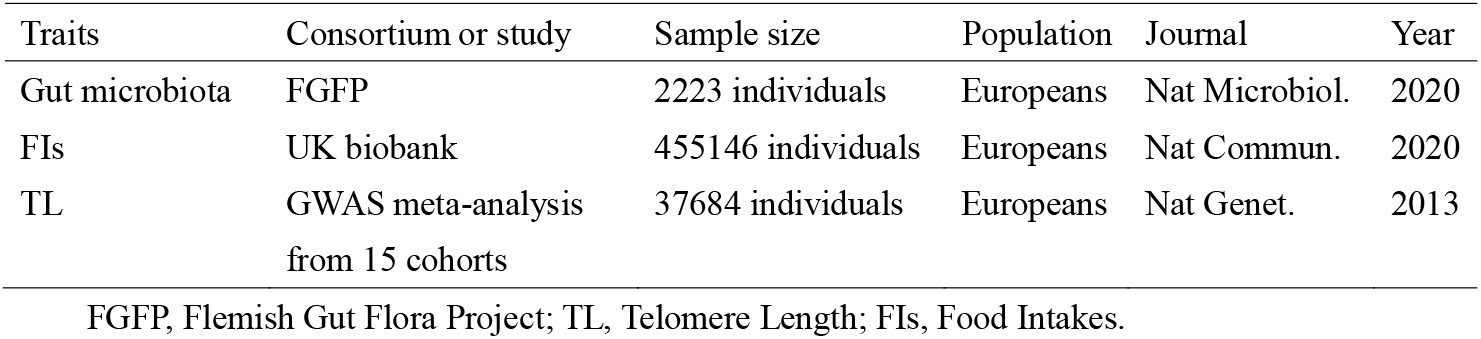
Description of GWAS summary data for gut microbiota, FIs and TL.

| Traits | Consortium or study | Sample size | Population | Journal | Year |
| --- | --- | --- | --- | --- | --- |
| Gut microbiota | FGFP | 2223 individuals | Europeans | Nat Microbiol. | 2020 |
| FIs | UK biobank | 455146 individuals | Europeans | Nat Commun. | 2020 |
| TL | GWAS meta-analysis<br>from 15 cohorts | 37684 individuals | Europeans | Nat Genet. | 2013 |

FGFP, Flemish Gut Flora Project; TL, Telomere Length; FIs, Food Intakes.

### 2.3 Statistical analysis

#### Calculation of significance threshold

To define our significance threshold for the two-sample MR analyses, we first run the HDL method to obtain the estimation of genetic correlations. 95 gut microbiota and 82 FIs are hierarchically clustered on the basis of genetic correlation values. Finally, eight independent gut microbiota clusters and eight independent FI-QTs clusters are obtained (Figure 2). Compared with LD Score Regression, HDL reduces the variance of genetic correlation estimations by about 60%, equivalent to a 2.5-fold increase in sample size. The efficiency improvement of HDL can be attributed to two reasons: (1) HDL uses more information on the relationship between test statistics and LD structure; (2) likelihood-based methods are more efficient than a method of moments [38]. Furthermore, another theoretical advantage of applying HDL on summary association statistics for binary phenotypes is that the HDL method models the GWAS test statistics whose distribution does not violate the normal assumption [38]. In order to derive the bi-directional causal relationships between FIs and gut microbial traits, we set our multiple-testing significance threshold at 7.81×10^−4^ (0.05/(8×8)). And the testing significance threshold is set at 6.25×10^−3^ (0.05/8) to obtain the causal relationships between FIs (or microbial traits) and TL.

**Figure 2.**
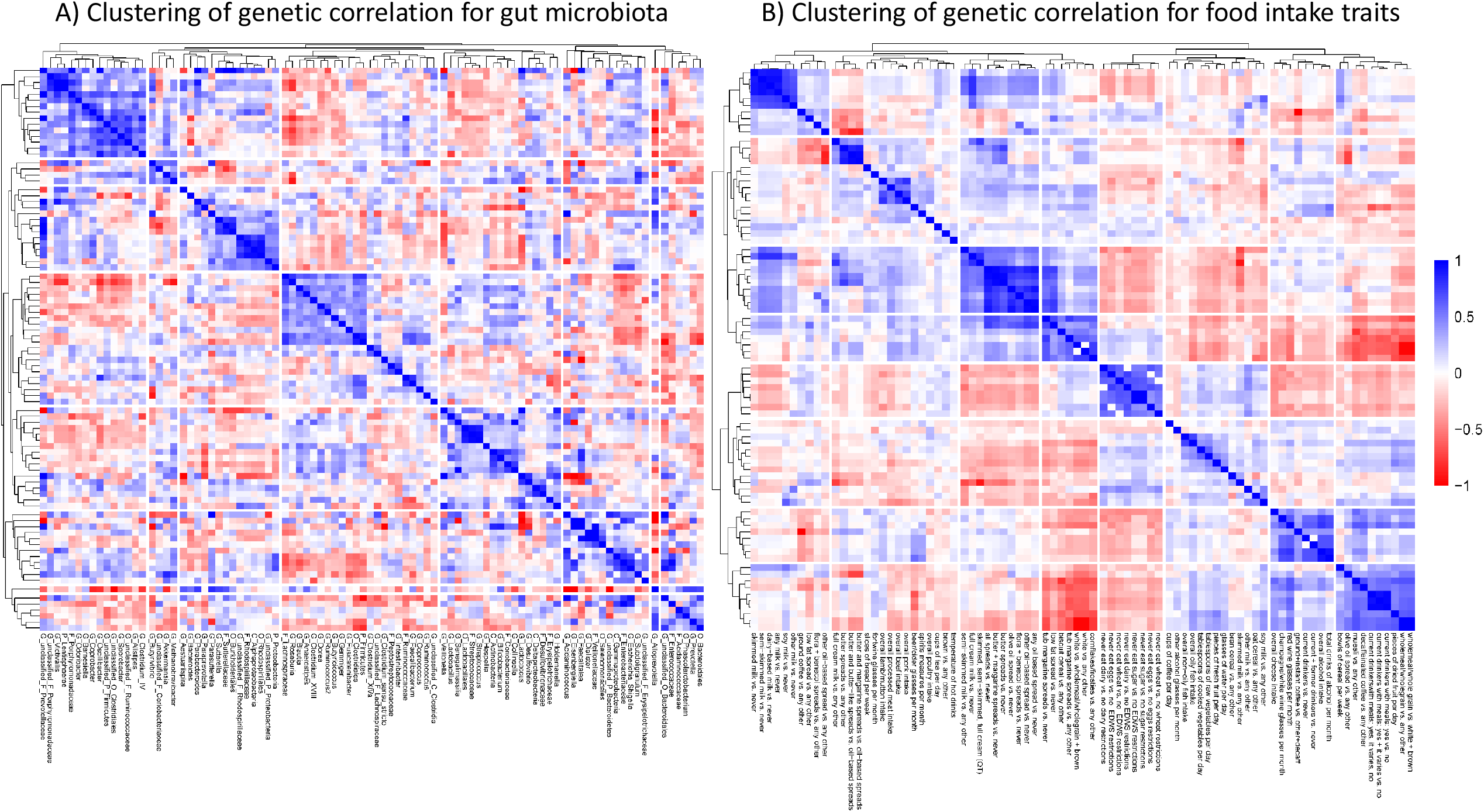
Clustering of genetic correlations for A) gut microbiota; and B) food intakes.

#### Mendelian Randomization

We perform bi-directional MR methods by traditional two-sample MR methods, including the inverse variance weighting method (IVW, fixed effects)[39], IVW (multiplicative random effects)[40], IVW radial [41], MR-Egger [42], weighted median[43], weighted mode[44], robust adjusted profile score (RAPS)[45]. The IVW (fixed effects) is the main analysis, for which SNPs need to satisfy the three core assumptions of MR. And other pleiotropy-robust methods are performed as auxiliary methods in our analysis. We also conduct several sensitivity analyses: Egger’s test [42] for detecting the pleiotropy, Wald ratio of each SNP for heterogeneity test[46] and Steiger test [47] for orienting the causal direction between imprecisely measured traits. The number of SNPs as IVs for each MR analysis is listed in the Supp Table 3-22. For each SNP, we calculate the F statistics and the traits variance of each SNP explained, which test the instrumental strength (Supp. Table 23-28).

##### GO enrichment analysis

To further explore the bio-function among gut microbiota, FIs and TL, we perform GO enrichment analysis based on lead SNPs for gut microbiota. Then, we map these SNPs of gut microbiota in different FIs or TL to the nearby genes. R packages used to implement the above statistical analyses include *HDL* and *TwosampleMR*, with R version 4.0.3. Statistical power is calculated in http://cnsgenomics.com/shiny/mRnd/.

## 3 Results

### 3.1 Bi-directional MR between FIs and gut microbiota

#### The causal effect of FIs on gut microbiota

Figure 3 A) shows the causal effect estimations of FIs on gut microbiota. Firstly, a positive association is found for the relationship of overall alcohol intake on Unclassified_K_Bacteria. Compared with any other coffee type, decaffeinated can causally increase the relative abundance of Acidaminococcaceae and ground can decrease it. The host-genetic-driven increase in Butyricicoccus is causally related to the intake of milk (including skimmed, semi-skimmed and full cream). In addition, compared with no eggs, dairy, wheat or sugar restrictions, never eating wheat has a positive association with Lentisphaerae and a negative association with Coprococcus. Furthermore, never eating wheat can reduce the relative abundance of Sutterella and increase the relative abundance of Victivallis and Lentisphaerae. Then, we also find suggestive association that the overall processed meat intake is causally related to the host-genetic-driven decrease in Desulfovibrionaceae. Eating more oil-based spread can causally increase the relative abundance of Lactococcus. Finally, intake more water per day is beneficial to the increase of the relative abundance of Barnesiella.

**Figure 3.**
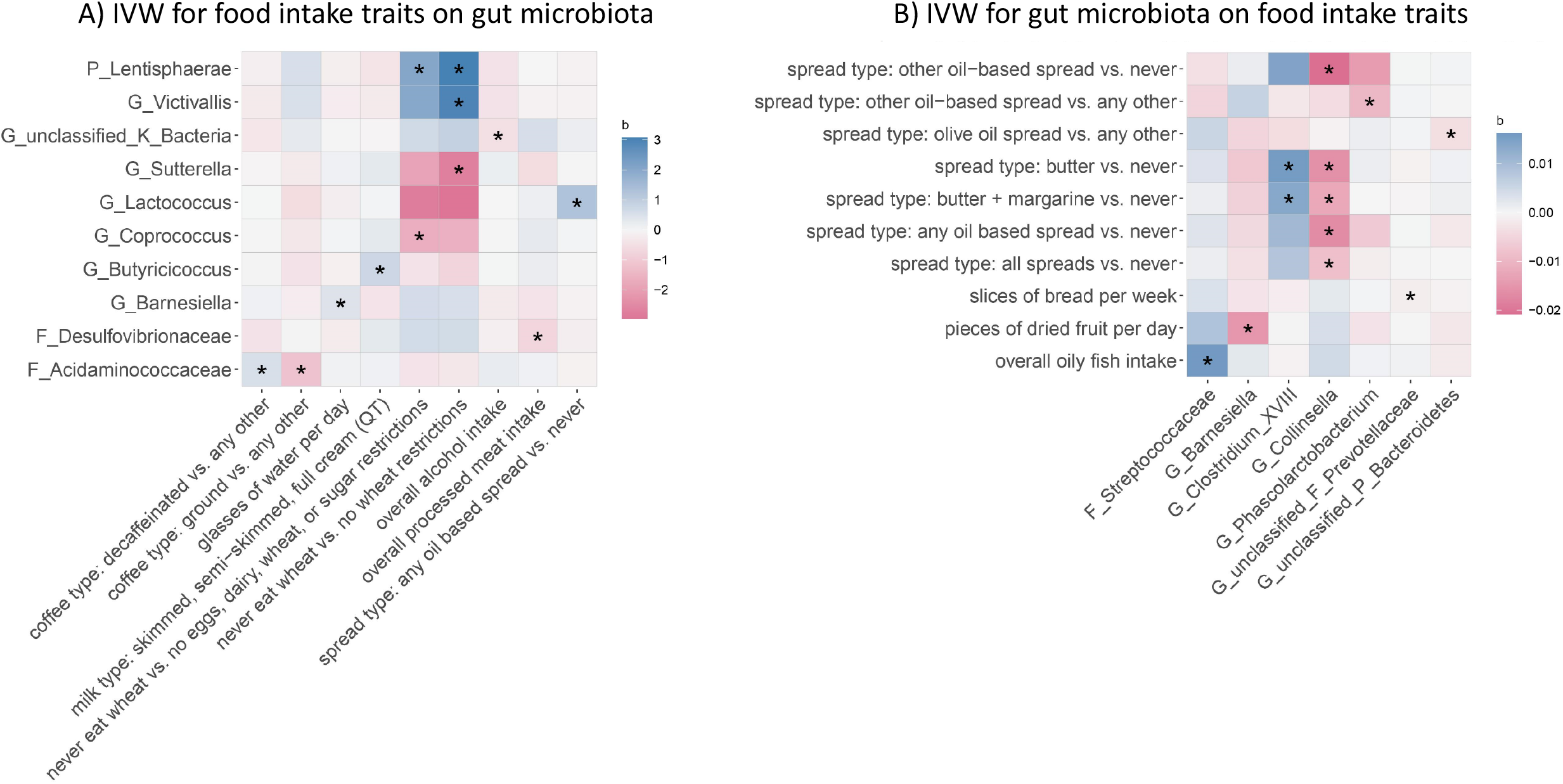
Causal effects estimation of bi-directional MR analysis between 82 gut microbiota and 95 food intakes. The cells marked with “*” represent the significant causal associations with Bonferroni correction P<0.05/64. The cell is colored according to the beta coefficients from inverse variance weighted (IVW) analysis. Other gut microbiota and food intakes which are not shown in the heatmap denote that there is no significant causal effect between them.

#### The causal effect of gut microbiota on FIs

In the opposite direction, the causal effect estimations of gut microbiota on FIs are shown in Figure 3 B). We find that host-genetic-driven increase in Unclassified_F_Prevotellaceae will lead to eat fewer slices of bread per week. Then, the opposite causal relationship of Barnesiella on dried fruit is also found. In addition, the intake of oily fish is positively affected by Streptococcaceae. In addition, the host-genetic-driven increase in Collinsella will causally decrease the intake of spread. However, the host-genetic-driven increase in Clostridium_XVIII can causally increase some intake of spread (including butter and margarine). Furthermore, the relative abundance of unclassified Bacteroidetes and Phascolarctobacterium negatively affect the intake of oil spread.

### 3.2 Bi-directional MR between TL and FIs

Figure 4 A) illustrates that a standard deviation increase in genetically influenced TL enhance the preference of alcohol, cereal (bran cereal) and coffee (ground and instant), while decreasing the preference of two types of cereal (cornflakes and frosties) and coffee (ground). If we relax the threshold at 0.05, the increase in TL also increases the intake of overall alcohol, decaffeinated coffee and red wine, and decreases the intake of poultry.

**Figure 4.**
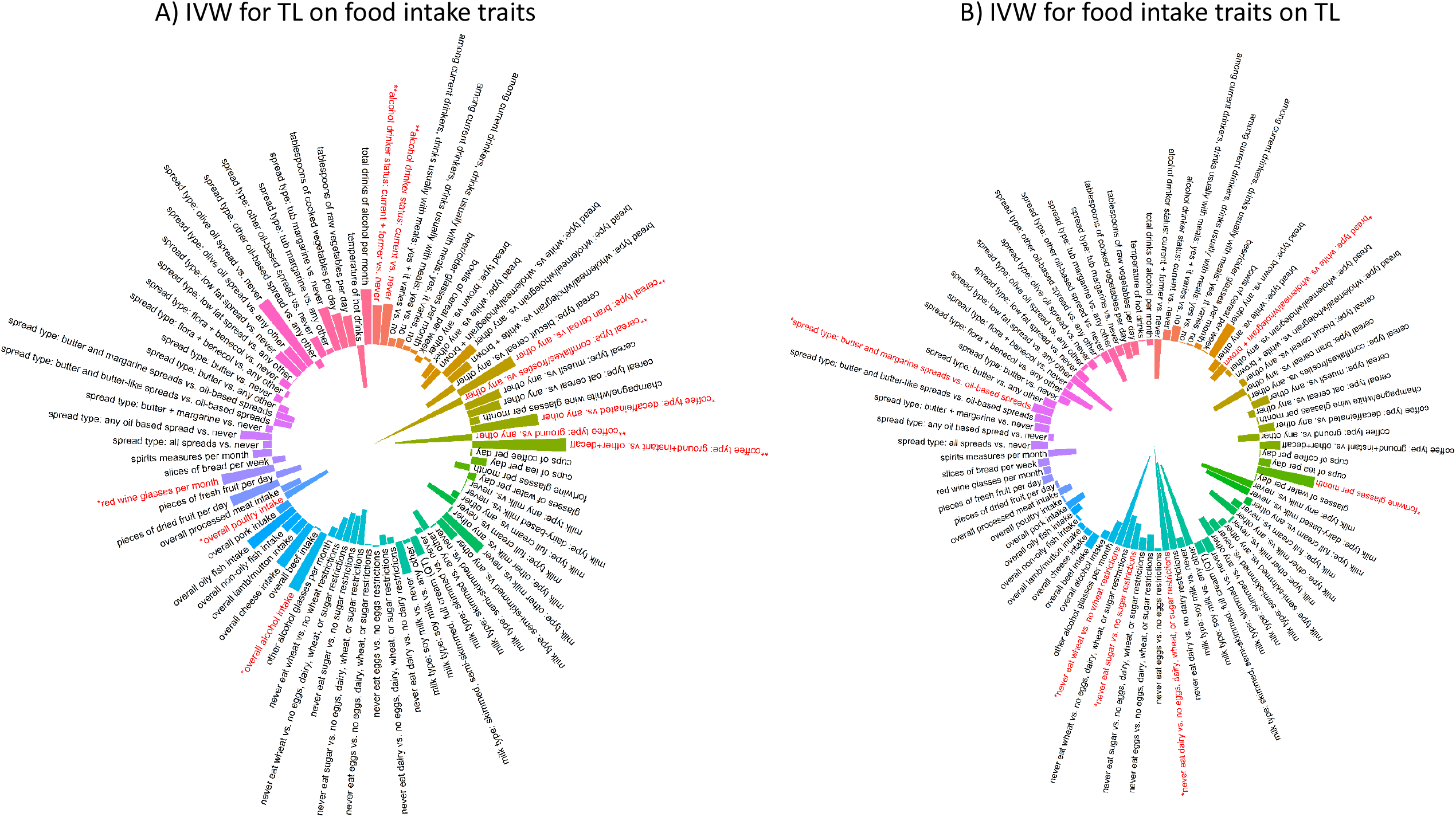
Causal effects estimation of bi-directional MR analysis between telomere length (TL) and 95 food intakes. The polar bars indicate the beta coefficients from inverse variance weighted (IVW) analysis. The bars marked with “**” represent the significant causal associations with Bonferroni correction P<0.05/8, while “*” represent the significant causal associations without any correction P<0.05.

For the opposite direction, we don’t find any causal relationships from FIs on the TL at the significant threshold value 6.25×10^−3^. If we relax the threshold at 0.05, we find that drinking fortwine and eating butter and margarine spread can increase the length of telomere. Inversely, eating white bread or never eating wheat, sugar or dairy can decrease the TL (Figure 4 B).

### 3.3 Bi-directional MR between TL and gut microbiota

Figure 5 A) shows that longer TL is causally related to the high relative abundance of G_Sutterella. We don’t find any other causal relationship between TL and gut microbiota at the significant threshold value 6.25×10^−3^. Thus, we relax the threshold at 0.05. Increased TL due to germline genetic variation was causally associated with higher relative abundance of Sutterellaceae, Sutterella, Veillonellaceae, Coprobacter and unclassified Clostridia and lower relative abundance of Gammaproteobacteria, Enterobacteriaceae, Anaerostipes, and Escherichia Shigella. For the opposite direction (Figure 5 B), we find that the host-genetic-driven decrease in Senegalimassilia taxa and unclassified Ruminococcaceae are positively related to the TL. On the contrary, host-genetic-driven decrease in Fusicatenibacter taxa and Sporobacter taxon is causally related to the TL.

**Figure 5.**
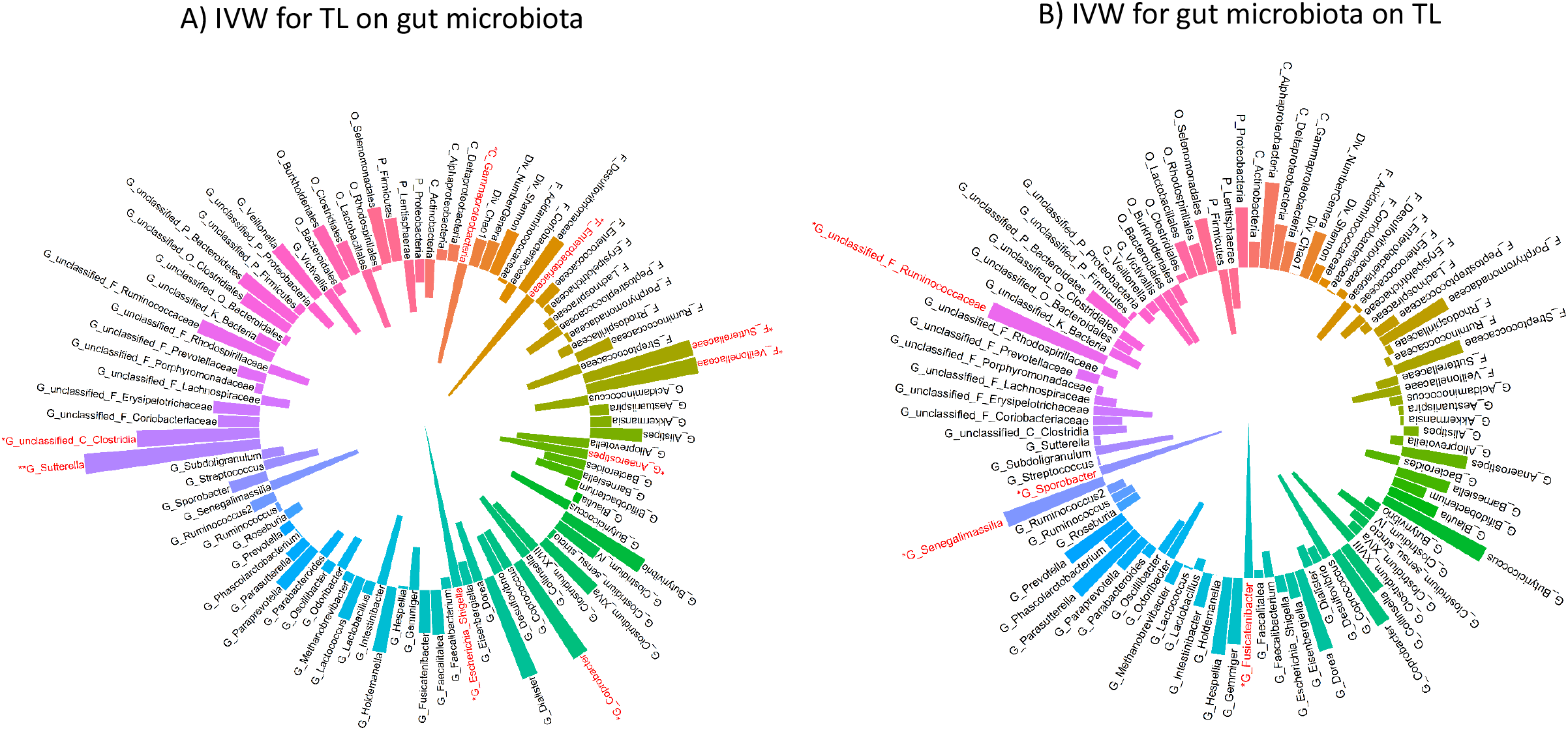
Causal effects estimation of bi-directional MR analysis between telomere length (TL) and 82 gut microbiota. The polar bar plot indicates the beta coefficients from inverse variance weighted (IVW) analysis. The bars marked with “**” represent the significant causal associations with Bonferroni correction P<0.05/8, while “*” represent the significant causal associations without any correction P<0.05.

There is 100% power to detect all the non-zero causal associations and this is strong enough for the causal effect among gut microbiota, FIs and TL, which are our main findings in the present study. Results of sensitivity analyses are listed in the Supplementary Notes (Figure S1 – S6) and Supplementary Table 29-34.

### 3.4 The network structure among FIs, gut microbiota and TL

The MR results-based summary network structure is shown in Figure 6. We find three main pathways among FIs, gut microbiota and TL. Firstly, the increased intake of water leads to the high relative abundance of Barnesiella, and further reduces the dried fruit intake. In addition, decreased relative abundance of Collinsella is beneficial to the intake of oil-based spread and further increases the relative abundance of Lactonccus. Furthermore, the TL affects the intake of ground coffee, and further influences the relative abundance of Acidaminococcaceae, which is also affected by decaffeinated coffee intake.

**Figure 6.**
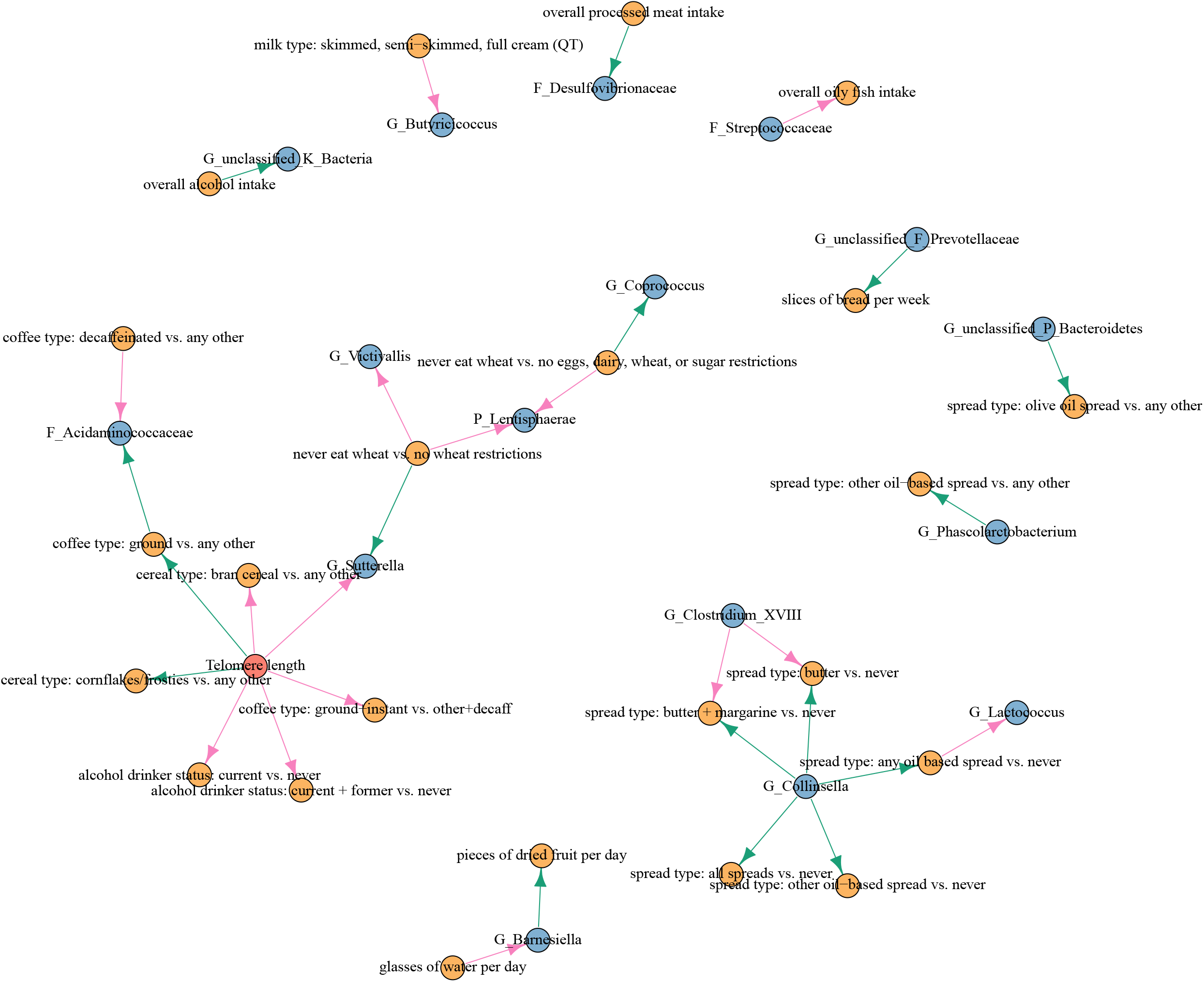
Network of gut microbiota, food intakes and telomere length (TL). The edge link two variables means there is a causal relationship between them after Bonferroni correction. Blue nodes represent gut microbiota, yellow nodes represent food intakes and red node represents telomere length. Pink edges represent positive causal effects while green edges represent negative causal effects.

### 3.5 GO enrichment analysis

For dried fruit intake per day and G_Barnesiella, 10 GO biological processes (e.g., dendrite morphogenesis, intrinsic component of synaptic membrane, and negative regulation of neuron projection development) were observed to be involved in both traits. We also found that 8 GO biological processes (regulation of GTPase activity, myosin V binding and startle response) were observed to be involved in slices of bread per week and G_unclassified_F_Prevotellaceae. For water intake and G_Barnesiella, 34 GO biological processes (e.g., positive regulation of synapse assembly, integral component of postsynaptic specialization membrane, and presynaptic endocytosis) were observed to be involved in both traits. For overall alcohol intake and G_unclassified_K_Bacteria, 38 GO biological processes (axonogenesis, axon development and axon guidance) were observed to be participants in both traits. In addition, 25 GO biological processes (regulation of GTPase activity, myosin V binding and startle response) were observed to be involved in overall processed meat intake and F_Desulfovibrionaceae. For telomere length and G_Sutterella, 13 GO biological processes (e.g., positive regulation of synapse assembly, integral component of postsynaptic specialization membrane, and presynaptic endocytosis) were observed to be involved in both traits. Results of GO enrichment analysis are listed in the Supplementary Notes (Figure S7 – S18).

## 4 Discussion

In this study, we performed a large-scale bi-directional MR study to explore the causal pathways among FIs, gut microbiota and TL. Overall, we found suggestive evidence of three main causal pathways among them. Eating one type of food (glasses of water per day) is able to affect the habit of eating another type of food (dried fruit intake) through its impact on the host gut microbiota (Barnesiella). The change of one gut microbiota taxon (Collinsella) in the host can causally influence the another gut microbiota taxon (Lactonccus) by affecting the diet habits (intake of oil-based based spread). The TL alters the dietary habits (intake of ground coffee) and further affects the gut microbiota (Acidaminococcaceae).

We found three main pathways from gut microbiota to TL through FIs. Some literatures provided evidence to support our results. For the first pathway, the intake of more water increased the relative abundance of Barnesiella, and further reduced the dried fruit intake. Drinking disinfected water could remove or control disease-causing microorganisms [48]. A study revealed that drinking more water may be an important factor in shaping the human gut microbiota, such as Bacteroides, in the US and UK populations [49]. A negative association was found between dried fruit supplement and Bacteroides relative abundance in a placebo-controlled, double-blind, randomized clinical trial[50]. For the second pathway, decreased relative abundance of Collinsella was beneficial to the intake of oil-based based spread and further increased the relative abundance of Lactonccus. The diversity in dietary oils might lead to the gut microbiota dysbiosis not only by decreasing their richness and diversity, but also by changing the Firmicutes to Bacteroidetes ratio [51]. In addition, Martín-Peláez et al. found a change in the intestinal microbiota following the intake of enriched virgin olive oil in 10 hypercholesterolemic individual [52,53]. The analytical sequencing of the intestinal microbiota showed that the reduction of Firmicutes and Blautia had a positive correlation with interleukin 1-β, TNF-α which would decrease following the administration of flaxseed oil [52,54]. For the third pathway, the TL affected the intake of ground coffee, and further influenced the relative abundance of Acidaminococcaceae. Coffee was made up of a variety of ingredients, including caffeine, polyphenols and fiber, and each of them had the potential to alter the gut microbiota [55]. Previous studies had investigated the coffee consumption was associated with Firmicutes [55], which was consistent with our results. The reason for the association between coffee and gut microbiota might be that the several putative stimuli (such as caffeine, polyphenol and melanoidin) could potentially stimulate gut epithelial cells, further influenced gut microbiota [56,57].

Furthermore, GO enrichment analysis found that larger number of GO biologic processes play key role in relationship among gut microbiota, FIs and TL. For example, the G_Sutterella was enriched in the ‘negative regulation of amyloid-beta formation pathway’, ‘negative regulation of amyloid precursor protein catabolic process pathway’ and other pathways associated with amyloid which has been confirmed to be associated with TL [58]. Furthermore, the G_Barnesiella was enriched in the ‘postsynaptic density organization pathway’, ‘postsynaptic specialization organization’ and other pathways related to the synapses. And Ana Adan indicate that dehydration increases your levels of the stress hormone cortisol, further disruptthe brain synapses [59]. In addition, there were three pathways associated with axon (‘axonogenesis pathway’,’ axon development pathway’,’ axon guidance pathway’’) enrich in G_unclassified_K_Bacteria. Supriti S et al. showed that alcohol intake induced structural alterations in axons [60].

Strengths of the present study include the genetic correlation and bi-directional MR design and the use of summary-level data from thus far the largest and latest GWASs. Using the clustering of genetic correlations to calculate a moderate significance threshold, which is not as strict as traditional Bonferroni correction. This design generally avoids bias due to reverse causation and confounding to obtain accurate results under MR assumptions. In addition, consistent results from several sensitivity analyses including the use of the other eight MR pleiotropic-robust methods indicate the robustness of our findings. The F statistics and three sensitivity analyses ensure the validity of MR core assumptions. On the other hand, the limitations of the current work include the inability to generalize the findings to other populations, as the sample predominantly comprised white adults of European descent. Besides, our analysis provides three potential causal pathways among gut microbiota, FIs and TL, which requires detailed external validation. Finally, we use a limited number of SNPs as IVs for gut microbiota, thus we cannot exclude the influence of weak instrument bias, although all genetic instruments are associated with the exposure (F-statistic > 10).

In summary, our findings support three potential causal pathways among gut microbiota, FIs and TL. Further investigations in understanding the underlying mechanisms of gut microbiota, and diet nutrition in the development of human aging are required.

## Data Availability

All the GWAS summary data are publically available. GWAS summary data for gut microbiota in FGFP can be download at https://doi.org/10.5523/bris.22bqn399f9i432q56gt3wfhzlc. GWAS summary data for Food intake in UK biobank can be download at https://www.kp4cd.org/dataset_downloads/t2d. GWAS summary data for telomere length can be download at https://www.ebi.ac.uk/gwas/publications/23535734.

## Acknowledgements

None.

## Author contributions

HL and FX conceived the study. YG, XL, SW, FQ contributed to data collection. LH and YY contributed to the data analysis. LZ and CJ contributed to the sensitivity analyses. LH and YY contributed to the application and wrote the manuscript. YW, YY, QL, YH, YF, WY modified the manuscript. All authors reviewed and approved the final manuscript.

## Ethics approval and patient consent statement

Ethical approval was not sought, because this study involved analysis of publicly available summary-level data from GWASs, and no individual-level data were used.

## Reference

1. Falony G, Joossens M, Vieira-Silva S, et al. Population-Level Analysis of Gut Microbiome Variation. Science (American Association for the Advancement of Science) 2016;352(6285):560–64.

2. Strasser B, Wolters M, Weyh C, Krüger K, Ticinesi A. The Effects of Lifestyle and Diet On Gut Microbiota Composition, Inflammation and Muscle Performance in Our Aging Society. Nutrients 2021;13(6):2045.

3. Zhernakova A, Kurilshikov A, Bonder MJ, et al. Population-Based Metagenomics Analysis Reveals Markers for Gut Microbiome Composition and Diversity. Science (American Association for the Advancement of Science) 2016;352(6285):565–69.

4. Lyte M. Probiotics Function Mechanistically as Delivery Vehicles for Neuroactive Compounds: Microbial Endocrinology in the Design and Use of Probiotics. Bioessays 2011;33(8):574–81.

5. Mayer EA, Rhee SH, Pothoulakis C. Principles and Clinical Implications of the Brain-Gut-Enteric Microbiota Axis. Nature clinical practice. Gastroenterology & hepatology 2009;6(5):306–14.

6. Norris V, Molina F, Gewirtz AT. Hypothesis: Bacteria Control Host Appetites. J Bacteriol 2013;195(3):411–16.

7. Alcock J, Maley CC, Aktipis CA. Is Eating Behavior Manipulated by the Gastrointestinal Microbiota? Evolutionary Pressures and Potential Mechanisms. Bioessays 2014;36(10):940–49.

8. Shikany JM, Demmer RT, Johnson AJ, et al. Association of Dietary Patterns with the Gut Microbiota in Older, Community-Dwelling Men. The American journal of clinical nutrition 2019;110(4):1003–14.

9. Claesson MJ, Jeffery IB, Conde S, et al. Gut Microbiota Composition Correlates with Diet and Health in the Elderly. Nature 2012;488(7410):178–84.

10. Bolte LA, Vich Vila A, Imhann F, et al. Long-Term Dietary Patterns are Associated with Pro-Inflammatory and Anti-Inflammatory Features of the Gut Microbiome. Gut 2021;70(7):1287–98.

11. Breton J, Tennoune N, Lucas N, et al. Gut Commensal E. Coli Proteins Activate Host Satiety Pathways Following Nutrient-Induced Bacterial Growth. Cell Metab 2016;23(2):324–34.

12. Fetissov SO. Role of the Gut Microbiota in Host Appetite Control: Bacterial Growth to Animal Feeding Behaviour. Nature reviews. Endocrinology 2017;13(1):11–25.

13. Rosenbaum M, Knight R, Leibel RL. The Gut Microbiota in Human Energy Homeostasis and Obesity. Trends Endocrin Met 2015;26(9):493–501.

14. Fetissov SO, Déchelotte P. The New Link Between Gut–Brain Axis and Neuropsychiatric Disorders. Curr Opin Clin Nutr 2011;14(5):477–82.

15. Blackburn EH, Epel ES, Lin J. Human Telomere Biology: A Contributory and Interactive Factor in Aging, Disease Risks, and Protection. Science (American Association for the Advancement of Science) 2015;350(6265):1193–98.

16. Turnbaugh PJ, Hamady M, Egholm M, et al. A Core Gut Microbiome in Obese and Lean Twins. Nature 2009;457(7228):480–84.

17. Maeda T, Horiuchi T, Makino N. Chromosomal Terminal Methylation Status is Associated with Gut Microbiotic Alterations. Mol Cell Biochem 2020;476(1):157–63.

18. García-Calzón S, Zalba G, Ruiz-Canela M, et al. Dietary Inflammatory Index and Telomere Length in Subjects with a High Cardiovascular Disease Risk From the Predimed-Navarra Study: Cross-Sectional and Longitudinal Analyses Over 5 Y. The American Journal of Clinical Nutrition 2015;102(4):897–904.

19. O’Toole PW, Jeffery IB. Gut Microbiota and Aging. Science (American Association for the Advancement of Science) 2015;350(6265):1214–15.

20. Biagi E, Franceschi C, Rampelli S, et al. Gut Microbiota and Extreme Longevity. Curr Biol 2016;26(11):1480–85.

21. Valdes AM, Andrew T, Gardner JP, et al. Obesity, Cigarette Smoking, and Telomere Length in Women. The Lancet 2005;366(9486):662–64.

22. Rafie N, Golpour HS, Barak F, Safavi SM, Miraghajani M. Dietary Patterns, Food Groups and Telomere Length: A Systematic Review of Current Studies. Eur J Clin Nutr 2017;71(2):151–58.

23. Turpin W, Espin-Garcia O, Xu W, et al. Association of Host Genome with Intestinal Microbial Composition in a Large Healthy Cohort. Nat Genet 2016;48(11):1413–17.

24. Sanna S, Zuydam NRV, Mahajan A, et al. Causal Relationships Among the Gut Microbiome, Short-Chain Fatty Acids and Metabolic Diseases. Nat Genet 2019;51(4):600–05.

25. Goodrich JK, Davenport ER, Beaumont M, et al. Genetic Determinants of the Gut Microbiome in Uk Twins. Cell Host Microbe 2016;19(5):731–43.

26. Groot HE, van de Vegte YJ, Verweij N, et al. Human Genetic Determinants of the Gut Microbiome and their Associations with Health and Disease: A Phenome-Wide Association Study. Sci Rep-Uk 2020;10(1):14771.

27. Lopera-Maya EA, Kurilshikov A, van der Graaf A, et al. Effect of Host Genetics On the Gut Microbiome in 7,738 Participants of the Dutch Microbiome Project. Nat Genet 2022;54(2):143.

28. Rode L, Bojesen SE, Weischer M, Nordestgaard BG. High Tobacco Consumption is Causally Associated with Increased All-Cause Mortality in a General Population Sample of 55 568 Individuals, but Not with Short Telomeres: A Mendelian Randomization Study. Int J Epidemiol 2014;43(5):1473–83.

29. Haycock PC, Burgess S, Nounu A, et al. Association Between Telomere Length and Risk of Cancer and Non-Neoplastic Diseases: A Mendelian Randomization Study. Jama Oncol 2017;3(636-51.

30. Kuo CL, Pilling LC, Kuchel GA, Ferrucci L, Melzer D. Telomere Length and Aging-Related Outcomes in Humans: A Mendelian Randomization Study in 261,000 Older Participants. Aging Cell 2019;18(6):e13017.

31. Hughes DA, Bacigalupe R, Wang J, et al. Genome-Wide Associations of Human Gut Microbiome Variation and Implications for Causal Inference Analyses. Nat Microbiol 2020;5(9):1079–87.

32. Sanderson E, Windmeijer F. A Weak Instrument F-Test in Linear Iv Models with Multiple Endogenous Variables. J Econometrics 2016;190(2):212–21.

33. Sudlow C, Gallacher J, Allen N, et al. Uk Biobank: An Open Access Resource for Identifying the Causes of a Wide Range of Complex Diseases of Middle and Old Age. Plos Med 2015;12(3):e1001779.

34. Cole JB, Florez JC, Hirschhorn JN. Comprehensive Genomic Analysis of Dietary Habits in Uk Biobank Identifies Hundreds of Genetic Associations. Nat Commun 2020;11(1):1467.

35. Codd V, Nelson CP, Albrecht E, et al. Identification of Seven Loci Affecting Mean Telomere Length and their Association with Disease. Nat Genet 2013;45(4):422–27.

36. Cawthon RM. Telomere Length Measurement by a Novel Monochrome Multiplex Quantitative Pcr Method. Nucleic Acids Res 2009;37(3):e21.

37. Cawthon RM. Telomere Measurement by Quantitative Pcr. Nucleic Acids Res 2002;30(10):e47.

38. Ning Z, Pawitan Y, Shen X. High-Definition Likelihood Inference of Genetic Correlations Across Human Complex Traits. In; 2020:223.

39. Hou L, Li H, Si S, et al. Exploring the Causal Pathway From Bilirubin to Cvd and Diabetes in the Uk Biobank Cohort Study: Observational Findings and Mendelian Randomization Studies. Atherosclerosis 2021;320(112-21.

40. Burgess S, Butterworth A, Thompson SG. Mendelian Randomization Analysis with Multiple Genetic Variants Using Summarized Data. Genet Epidemiol 2013;37(7):658–65.

41. Bowden J, Spiller W, Del Greco M F, et al. Improving the Visualization, Interpretation and Analysis of Two-Sample Summary Data Mendelian Randomization Via the Radial Plot and Radial Regression. Int J Epidemiol 2018;47(6):2100.

42. Burgess S, Thompson SG. Interpreting Findings From Mendelian Randomization Using the Mr-Egger Method. Eur J Epidemiol 2017;32(5):377–89.

43. Bowden J, Davey Smith G, Haycock PC, Burgess S. Consistent Estimation in Mendelian Randomization with some Invalid Instruments Using a Weighted Median Estimator. Genet Epidemiol 2016;40(4):304–14.

44. Burgess S, Zuber V, Gkatzionis A, Foley CN. Modal-Based Estimation Via Heterogeneity-Penalized Weighting: Model Averaging for Consistent and Efficient Estimation in Mendelian Randomization When a Plurality of Candidate Instruments are Valid. Int J Epidemiol 2018;47(4):1242–54.

45. Zhao Q, Wang J, Hemani G, Bowden J, Small DS. Statistical Inference in Two-Sample Summary-Data Mendelian Randomization Using Robust Adjusted Profile Score. The Annals of statistics 2020;48(3):1742.

46. Burgess S, Bowden J, Fall T, Ingelsson E, Thompson SG. Sensitivity Analyses for Robust Causal Inference From Mendelian Randomization Analyses with Multiple Genetic Variants. Epidemiology 2017;28(1):30–42.

47. Lutz SM, Wu AC, Hokanson JE, Vansteelandt S, Lange C. Caution Against Examining the Role of Reverse Causality in Mendelian Randomization. Genet Epidemiol 2021;45(5):445–54.

48. Dai Z, Sevillano-Rivera MC, Calus ST, et al. Disinfection Exhibits Systematic Impacts On the Drinking Water Microbiome. Microbiome 2020;8(1):42.

49. Vanhaecke T, Bretin O, Poirel M, Tap J. Drinking Water Source and Intake are Associated with Distinct Gut Microbiota Signatures in Us and Uk Populations. The Journal of nutrition 2022;152(1):171–82.

50. van der Merwe M, Moore D, Hill JL, et al. The Impact of a Dried Fruit and Vegetable Supplement and Fiber Rich Shake On Gut and Health Parameters in Female Healthcare Workers: A Placebo-Controlled, Double-Blind, Randomized Clinical Trial. Microorganisms (Basel) 2021;9(4):843.

51. Ye Z, Xu Y, Liu Y. Influences of Dietary Oils and Fats, and the Accompanied Minor Content of Components On the Gut Microbiota and Gut Inflammation: A Review. Trends Food Sci Tech 2021;113(255-76.

52. Merra G, Noce A, Marrone G, et al. Influence of Mediterranean Diet On Human Gut Microbiota. Nutrients 2020;13(1):7.

53. Martín-Peláez S, Castañer O, Solà R, et al. Influence of Phenol-Enriched Olive Oils On Human Intestinal Immune Function. Nutrients 2016;8(4):213.

54. Zhu L, Sha L, Li K, et al. Dietary Flaxseed Oil Rich in Omega-3 Suppresses Severity of Type 2 Diabetes Mellitus Via Anti-Inflammation and Modulating Gut Microbiota in Rats. Lipids Health Dis 2020;19(1):20.

55. Diamond E, Hewlett K, Penumutchu S, Belenky A, Belenky P. Coffee Consumption Modulates Amoxicillin-Induced Dysbiosis in the Murine Gut Microbiome. Front Microbiol 2021;12(637282.

56. Nakayama T, Oishi K. Influence of Coffee (Coffea Arabica) and Galacto-Oligosaccharide Consumption On Intestinal Microbiota and the Host Responses. Fems Microbiol Lett 2013;343(2):161–68.

57. Daglia M, Papetti A, Grisoli P, et al. Isolation, Identification, and Quantification of Roasted Coffee Antibacterial Compounds. J Agr Food Chem 2007;55(25):10208–13.

58. Rolyan H, Scheffold A, Heinrich A, et al. Telomere Shortening Reduces Alzheimer’s Disease Amyloid Pathology in Mice. Brain 2011;134(Pt 7):2044–56.

59. Adan A. Cognitive Performance and Dehydration. J Am Coll Nutr 2012;31(2):71–78.

60. Samantaray S, Knaryan VH, Patel KS, et al. Chronic Intermittent Ethanol Induced Axon and Myelin Degeneration is Attenuated by Calpain Inhibition. Brain Res 2015;1622;7–21.

