## Supplemental Notes for "Identification of Causal Pathways among Gut Microbiota, Food Intake and Telomere Length: A Mendelian Randomization Study"

**Content**

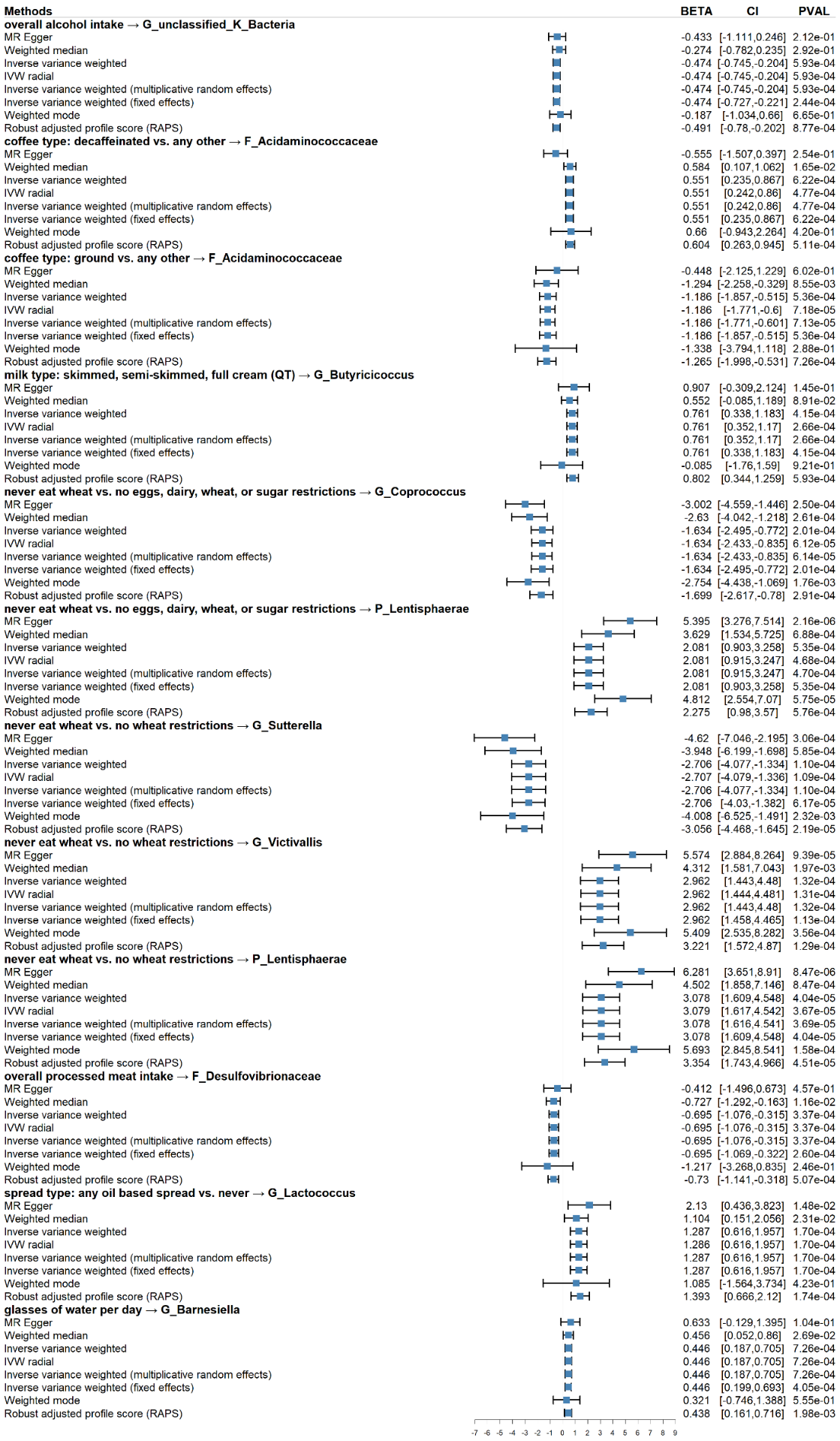

### Figure S1. Sensitive analysis of significant causal relationships of food intake on microbiome.

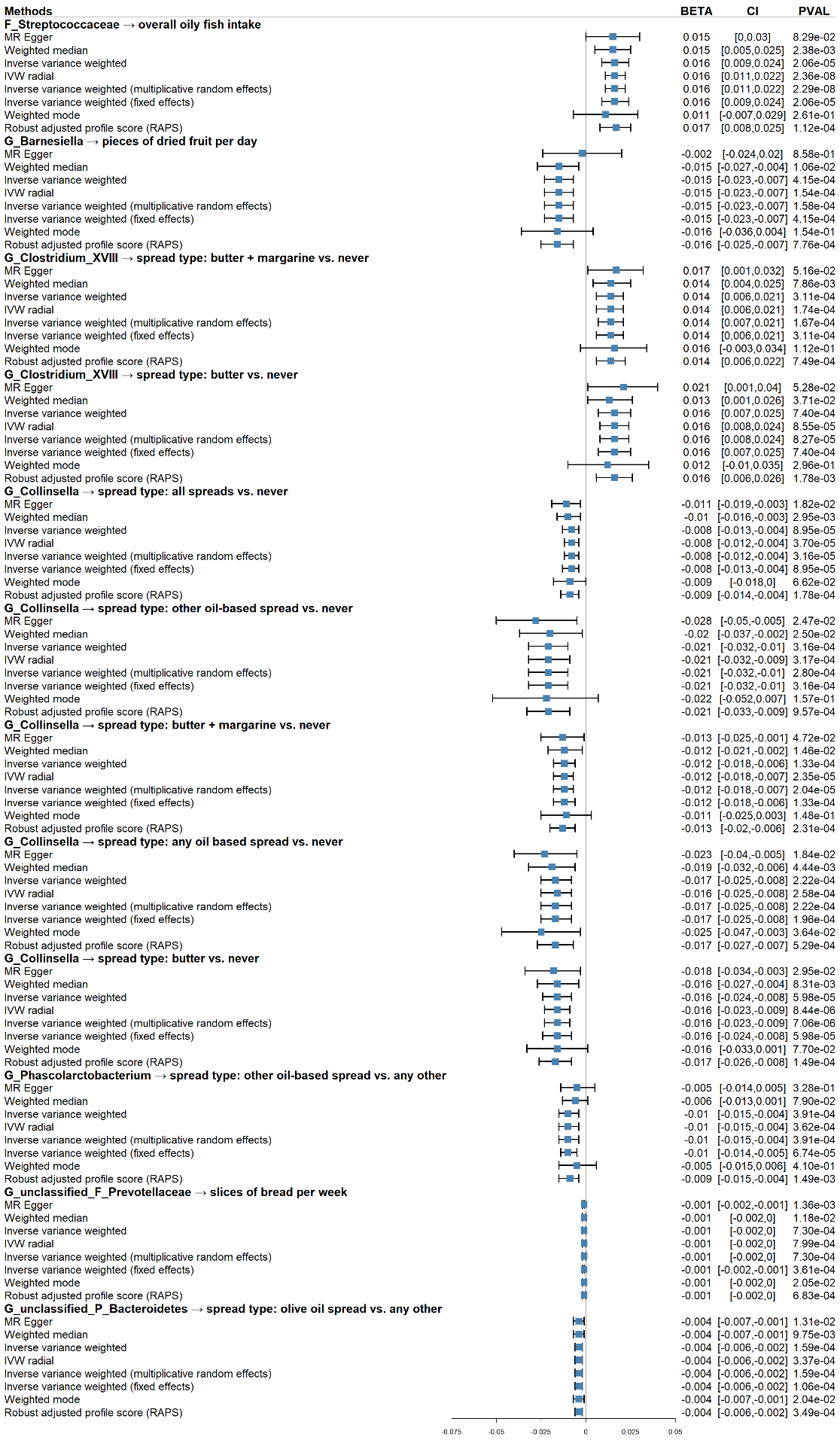

### Figure S2. Sensitive analysis of significant causal relationships of microbiome on food intake.

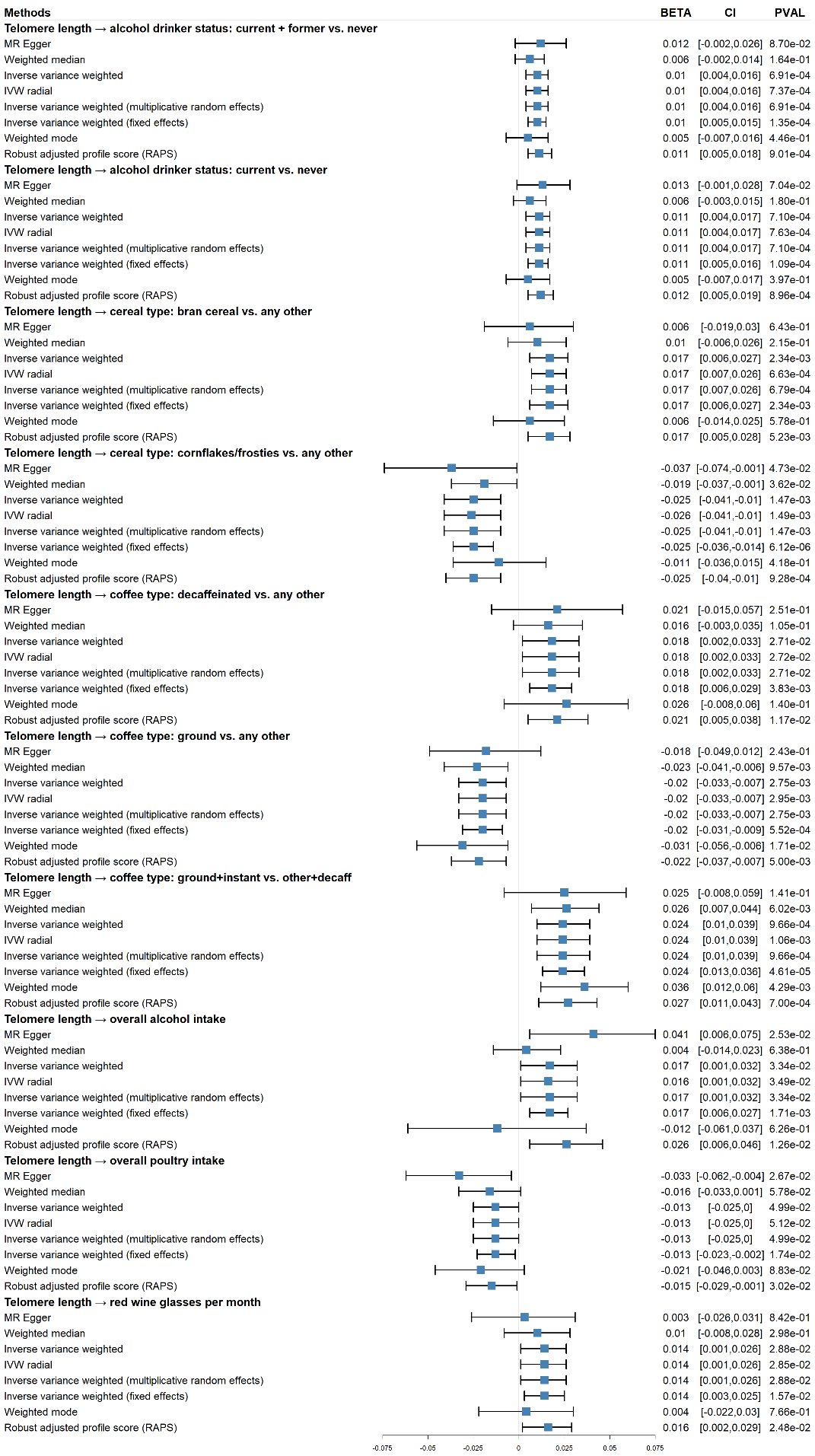

### Figure S3. Sensitive analysis of significant causal relationships of telomere length on food intake.

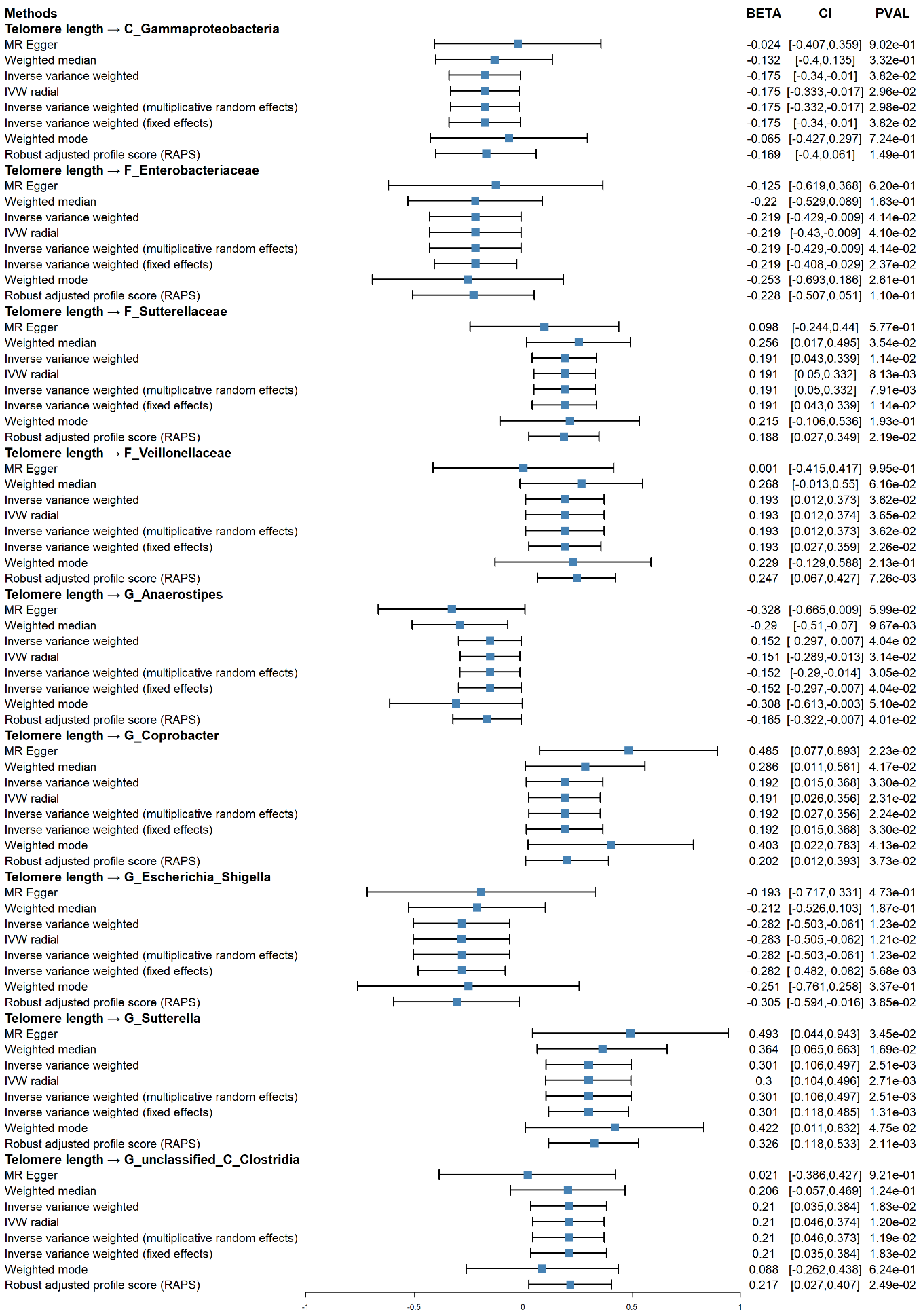

### Figure S4. Sensitive analysis of significant causal relationships of telomere length on microbiome.

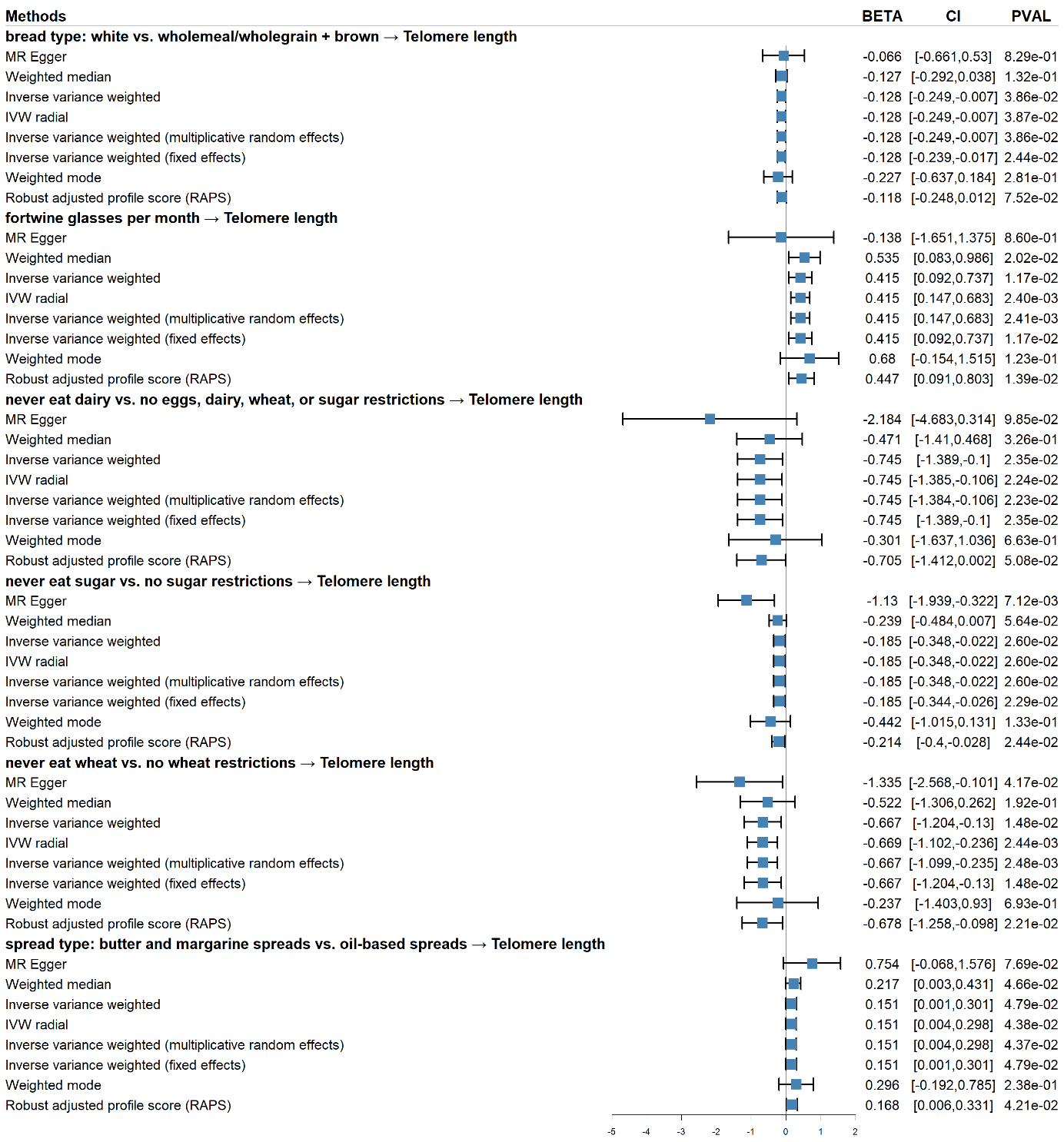

### Figure S5. Sensitive analysis of significant causal relationships of food intake on telomere length.

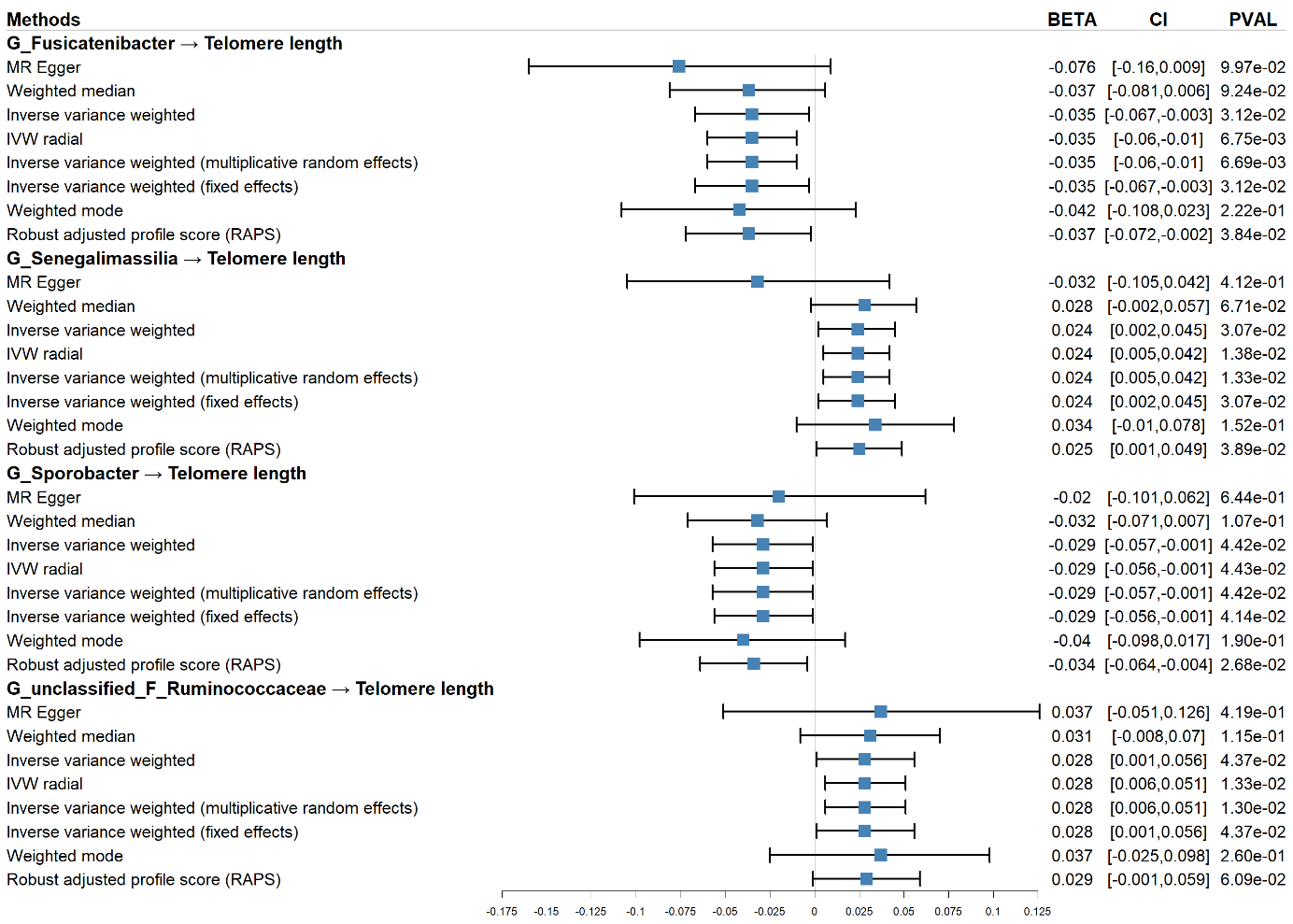

### Figure S6. Sensitive analysis of significant causal relationships of microbiome on telomere length.

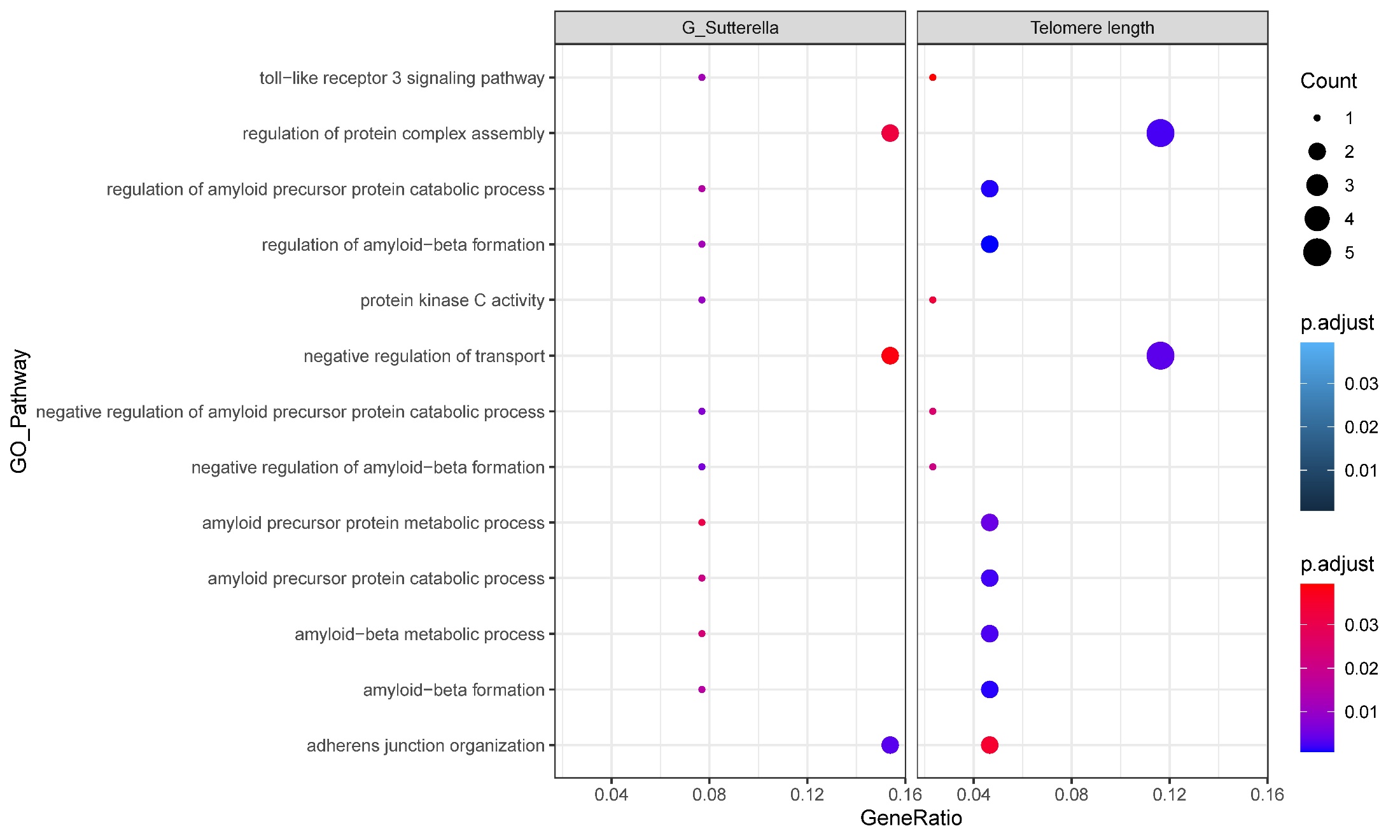

### Figure S7. GO enrichment analysis of telomere length and G_Sutterella (gene ratio and pvalue).

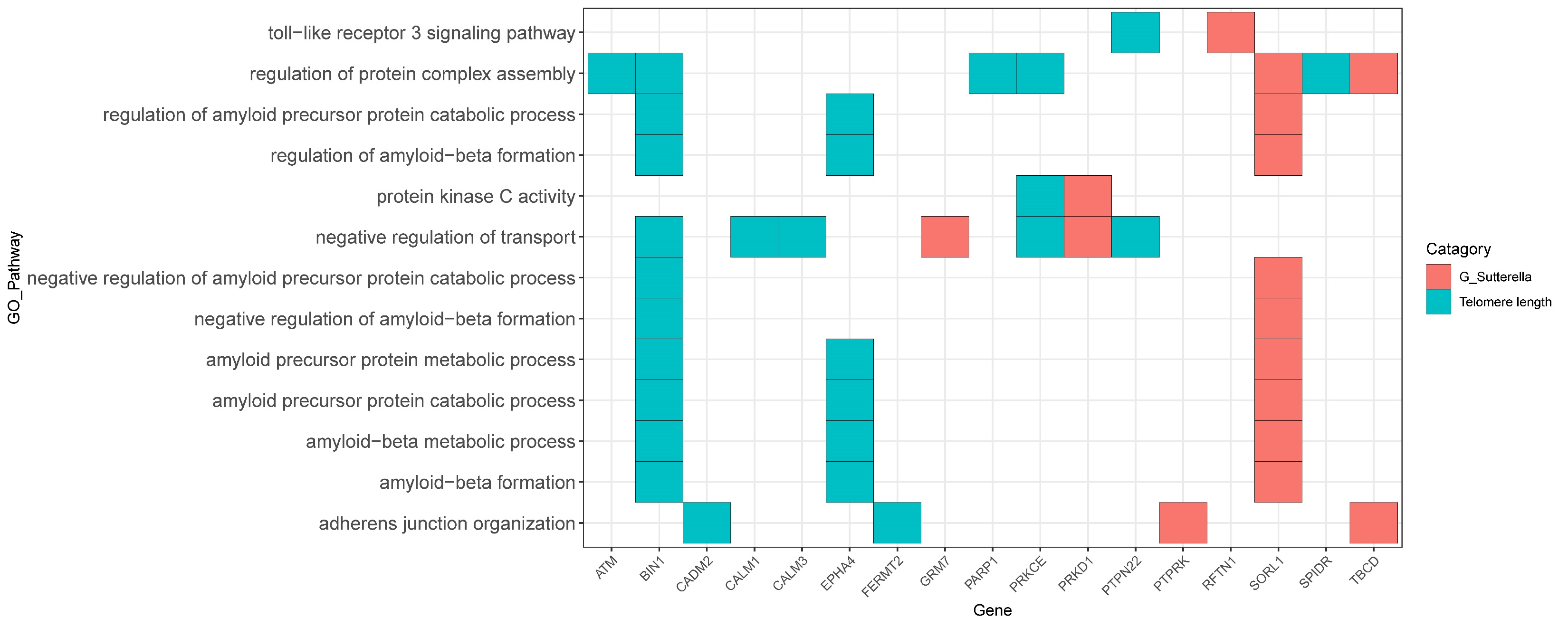

### Figure S8. GO enrichment analysis of telomere length and G_Sutterella (gene).

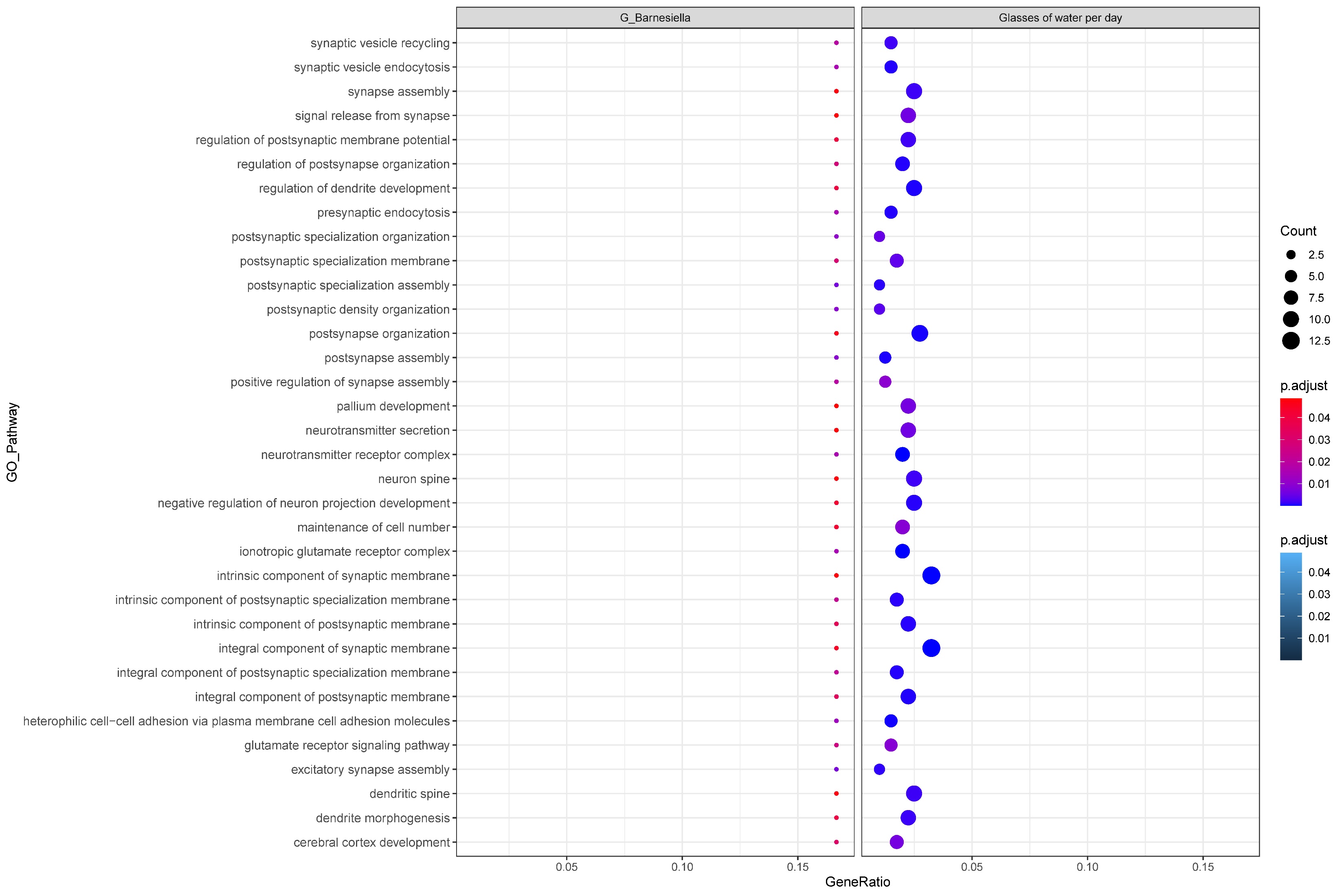

### Figure S9. GO enrichment analysis of glasses of water per day and G_Barnesiella (gene ratio and pvalue).

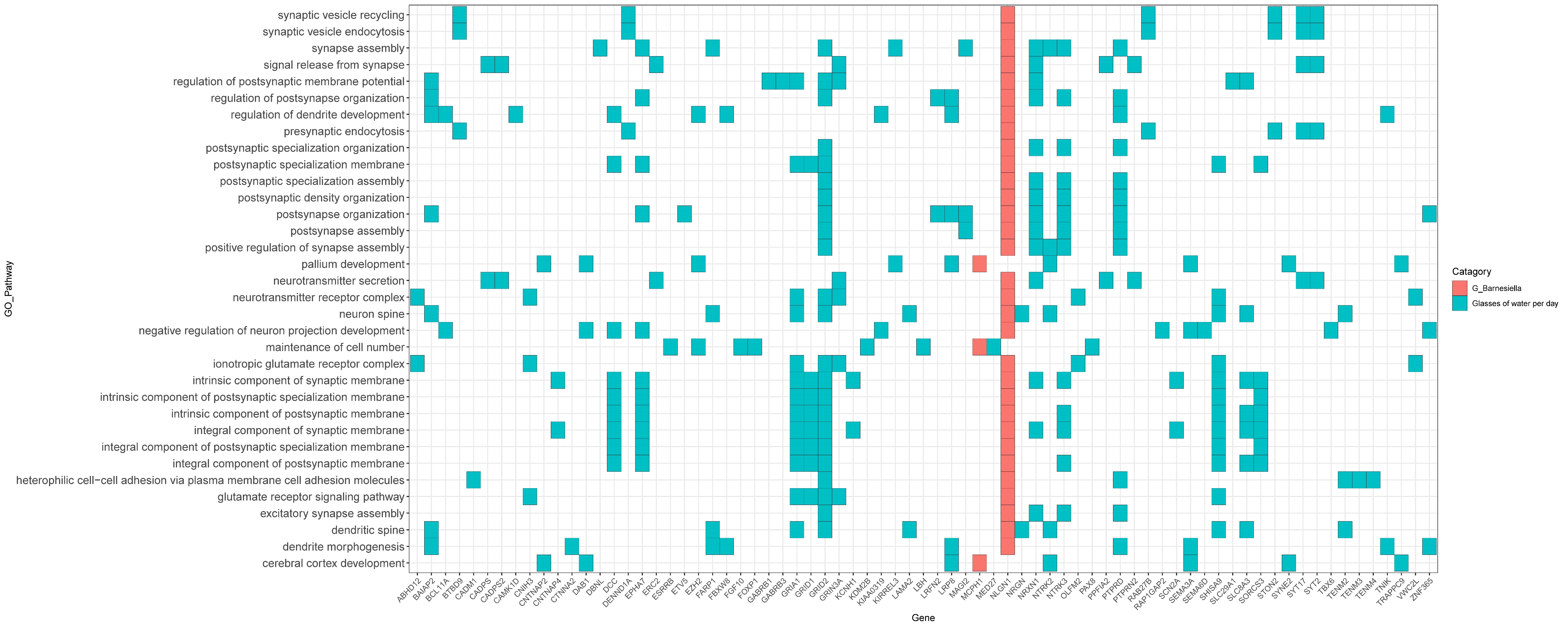

### Figure S10. GO enrichment analysis of glasses of water per day and G_Barnesiella (gene).

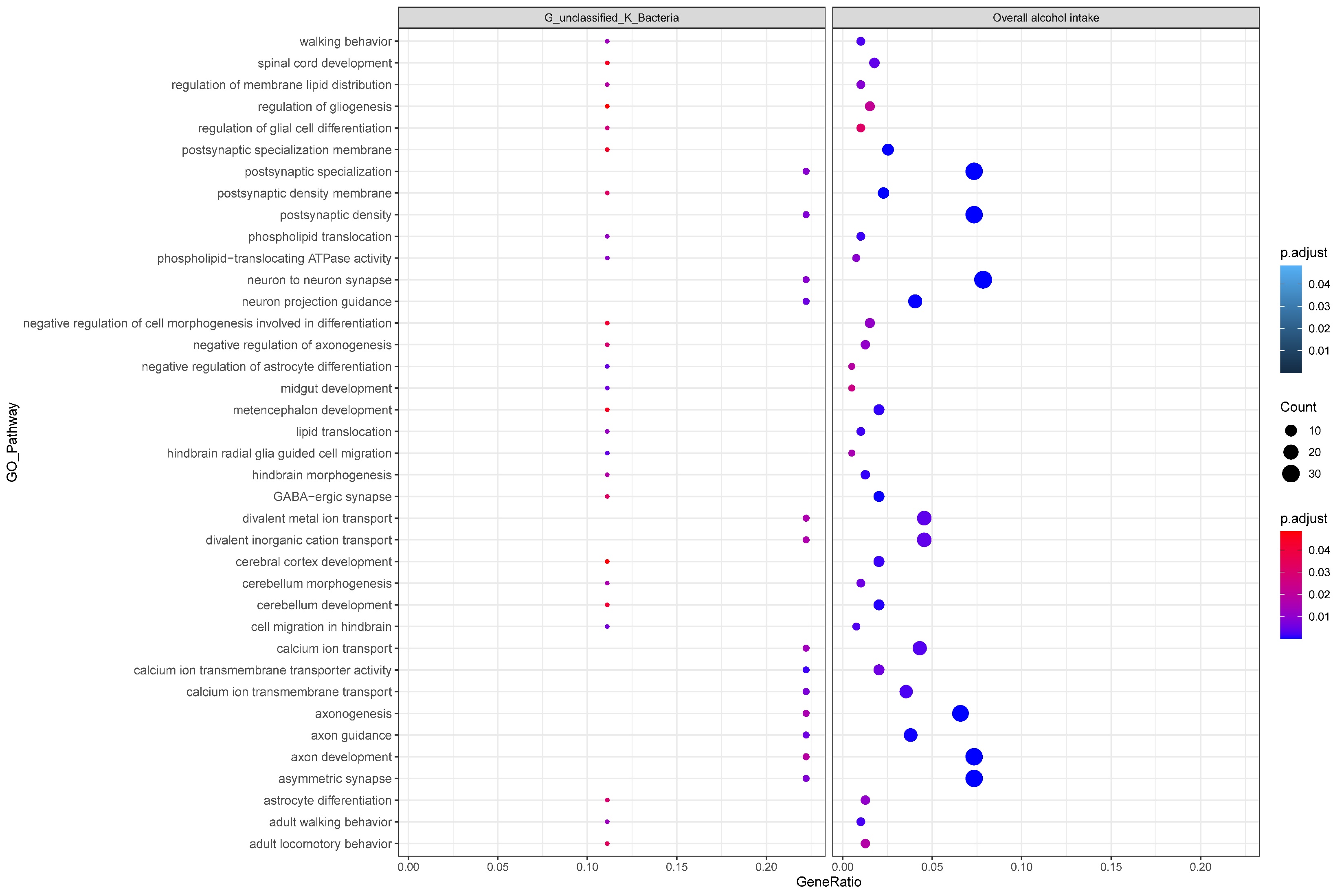

### Figure S11. GO enrichment analysis of overall alcohol intake and G_unclasssified_K_Bacteria (gene ratio and pvalue).

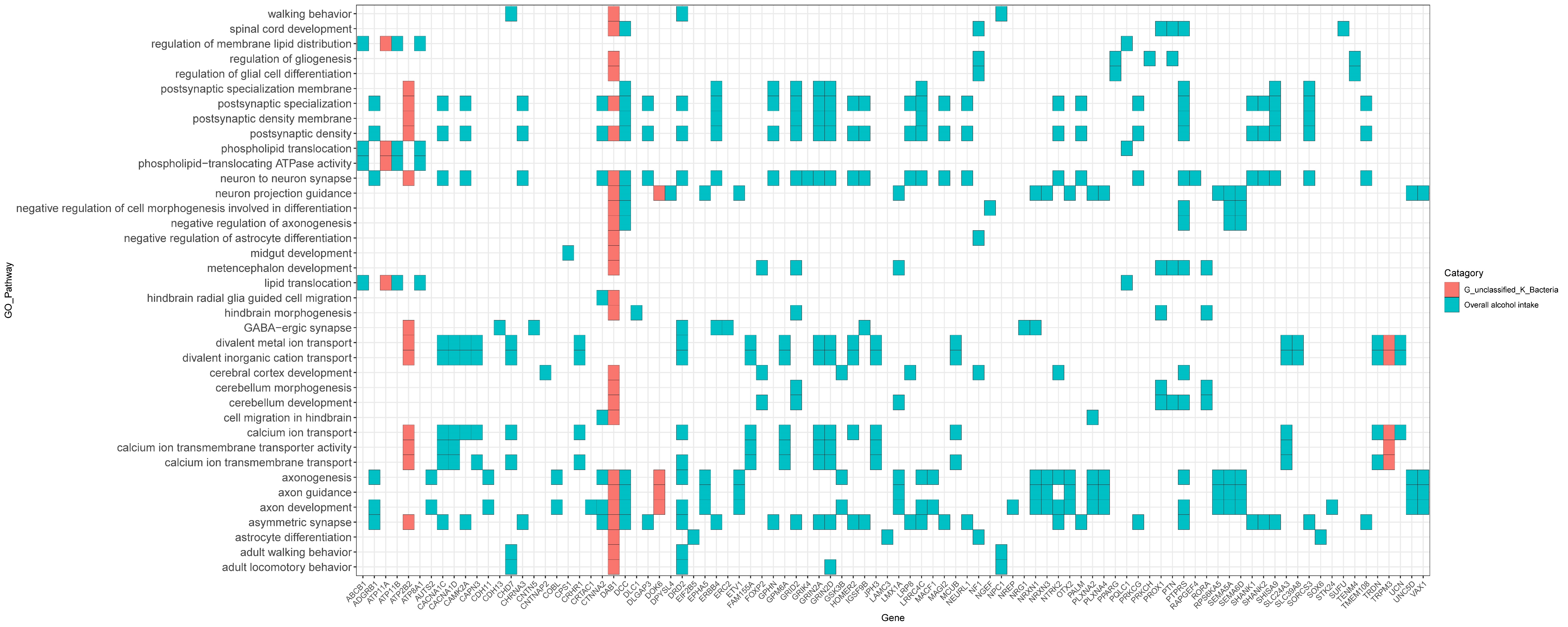

### Figure S12. GO enrichment analysis of overall alcohol intake and G_unclasssified_K_Bacteria (gene).

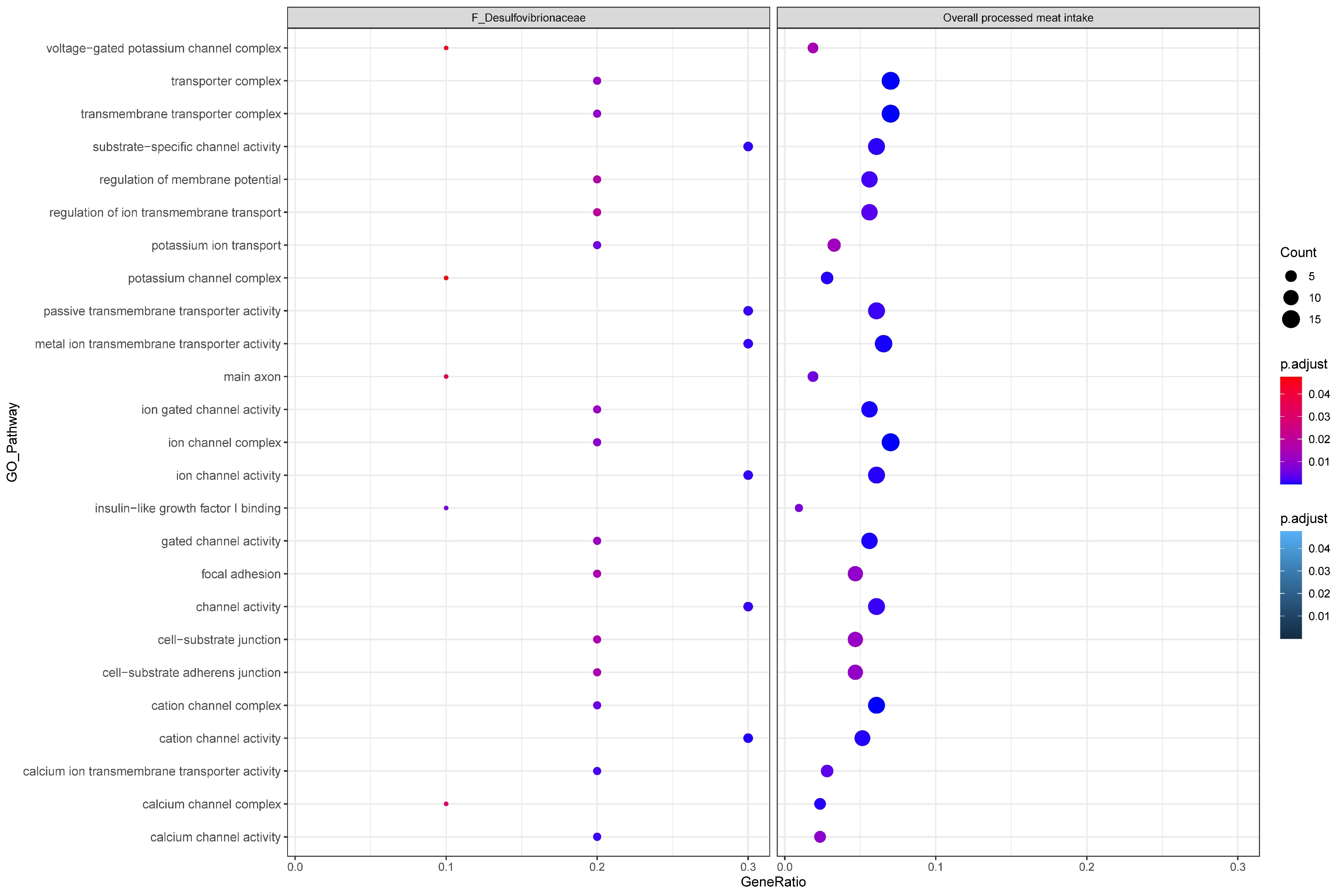

### Figure S13. GO enrichment analysis of overall processed meant intake and F_Desulfovibrionaceae (gene ratio and pvalue).

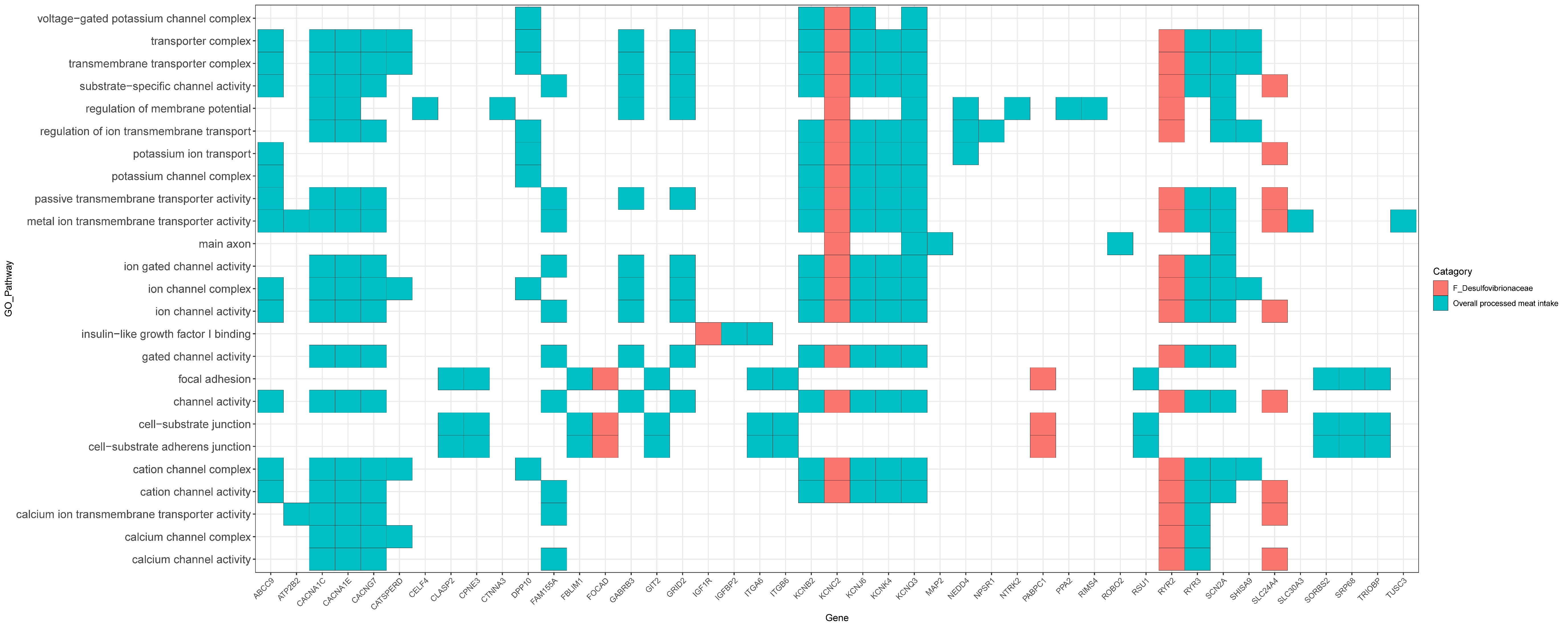

### Figure S14. GO enrichment analysis of overall processed meant intake and F_Desulfovibrionaceae (gene).

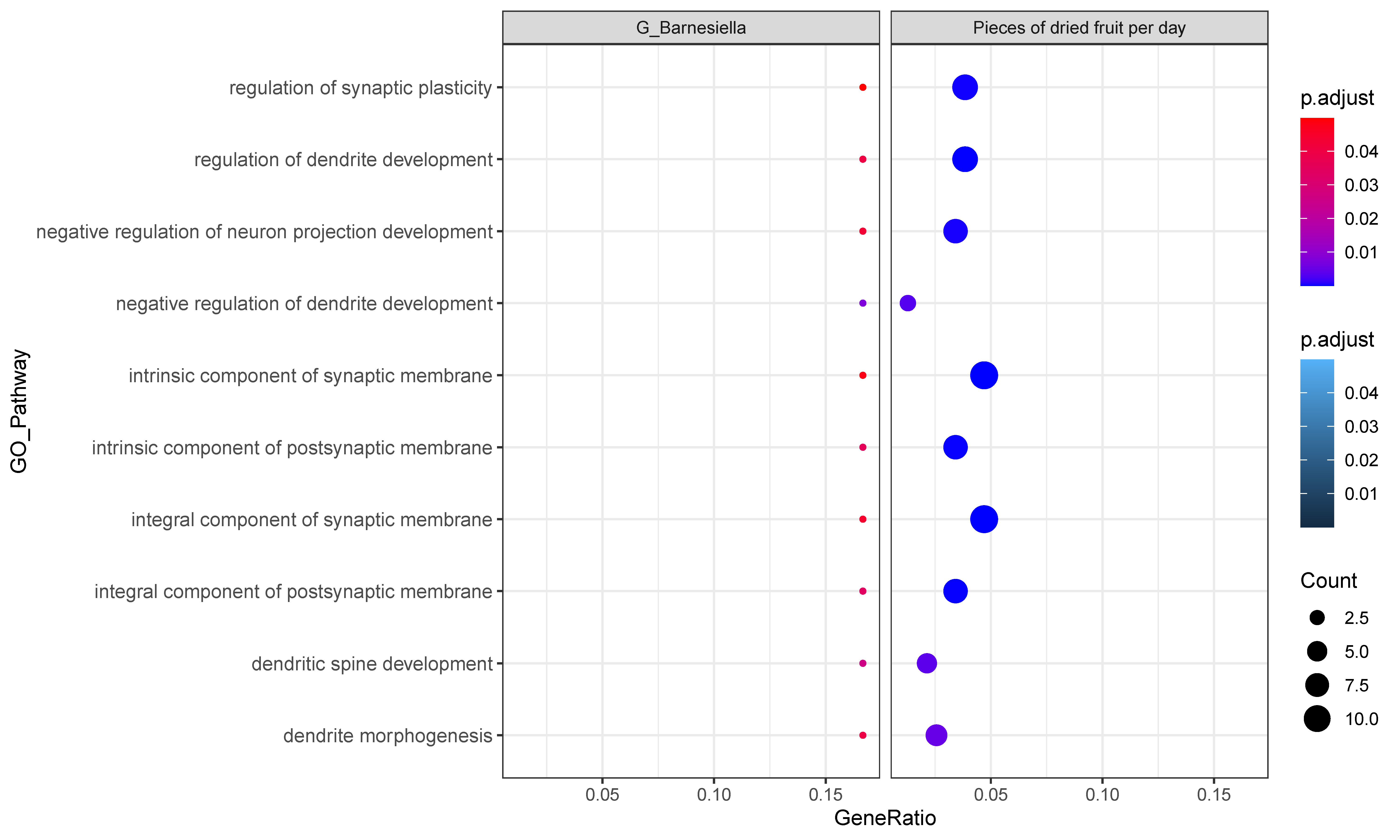

### Figure S15. GO enrichment analysis of overall processed meant intake and F_Desulfovibrionaceae (gene ratio and pvalue).

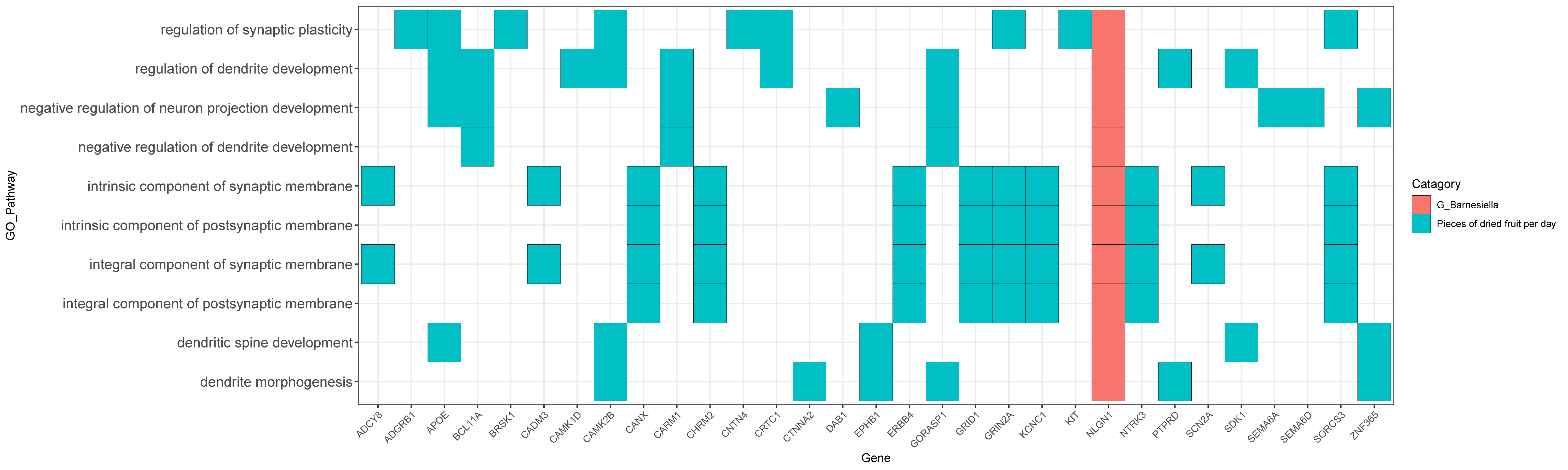

### Figure S16. GO enrichment analysis of pieces of dries fruit per day and G_Barnesiella (gene).

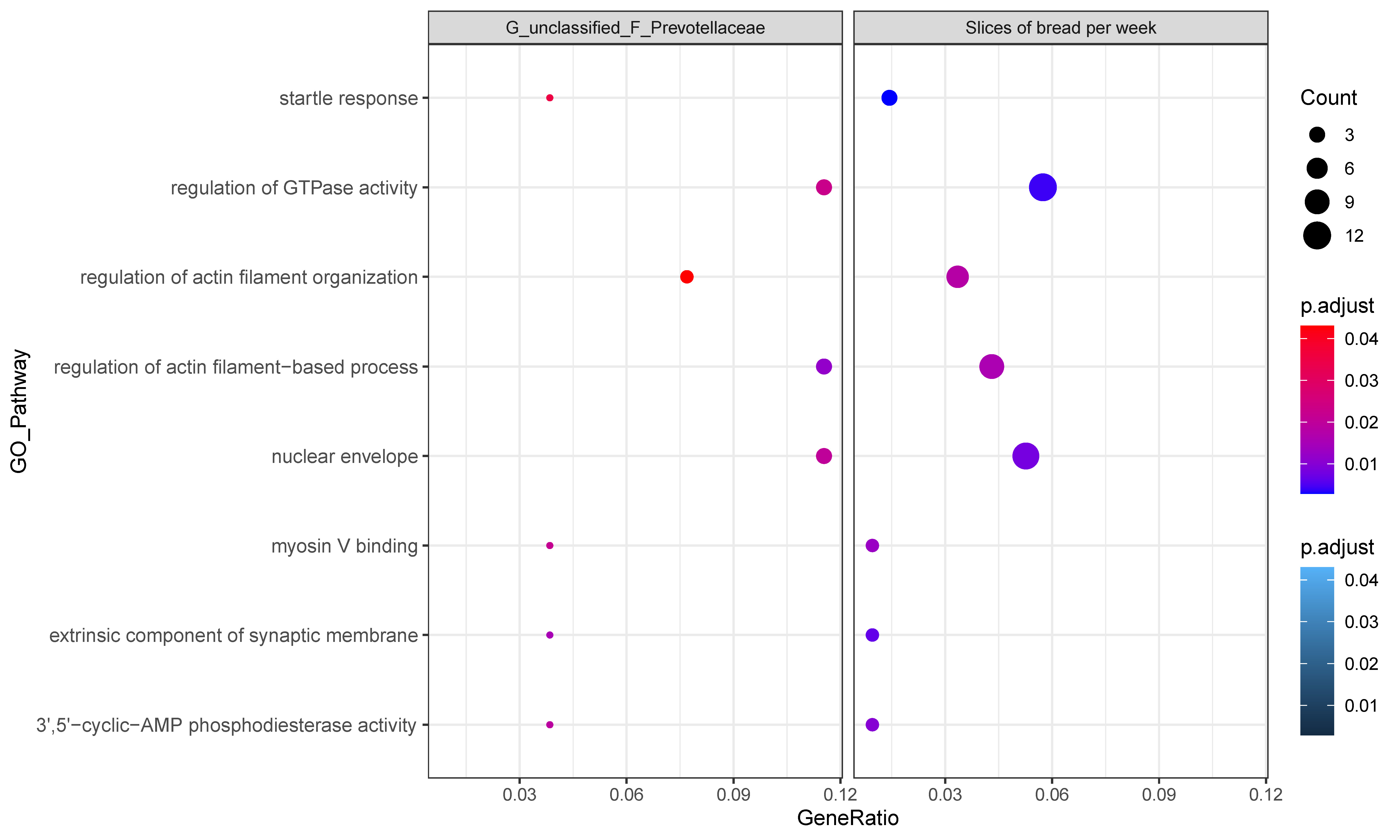

### Figure S17. GO enrichment analysis of slices of bread per week and G_unclassified_F_Prevotellaceae (gene ratio and pvalue).

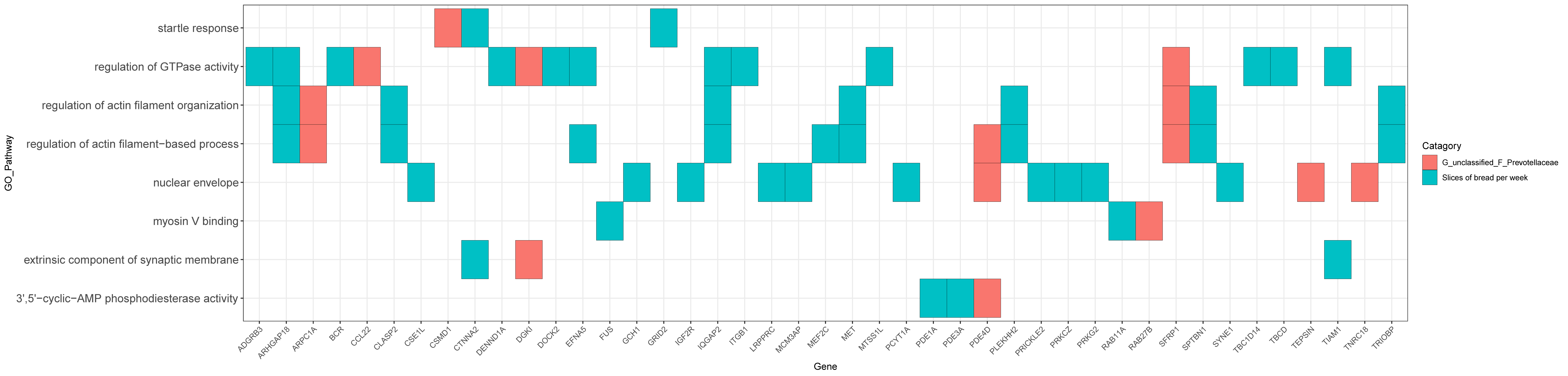

### Figure S18. GO enrichment analysis of slices of bread per week and G_unclassified_F_Prevotellaceae (gene).
